# Grey and white matter micro-structure is associated with polygenic risk for schizophrenia

**DOI:** 10.1101/2021.02.06.21251073

**Authors:** Eva-Maria Stauffer, Richard A.I. Bethlehem, Varun Warrier, Graham K. Murray, Rafael Romero-Garcia, Jakob Seidlitz, Edward T. Bullmore

## Abstract

**Background:** Recent discovery of approximately 270 common genetic variants associated with schizophrenia has enabled polygenic risk scores (PRS) to be measured in the population. We hypothesized that normal variation in PRS would be associated with magnetic resonance imaging (MRI) phenotypes of brain morphometry and tissue composition.

**Methods:** We used the largest extant genome-wide association dataset (N = 69,369 cases and N = 236,642 healthy controls) to measure PRS for schizophrenia in a large sample of adults from the UK Biobank (Nmax = 29,878) who had multiple micro- and macro-structural MRI metrics measured at each of 180 cortical areas, seven subcortical structures, and 15 major white matter tracts. Linear mixed effect models were used to investigate associations between PRS and brain structure at global and regional scales, controlled for multiple comparisons.

**Results:** Polygenic risk was significantly associated with reduced neurite density index (NDI) at global brain scale, at 149 cortical regions, five subcortical structures and 14 white matter tracts. Other micro-structural parameters, e.g., fractional anisotropy, that were correlated with NDI were also significantly associated with PRS. Genetic effects on multiple MRI phenotypes were co-located in temporal, cingulate and prefrontal cortical areas, insula, and hippocampus. Post-hoc bidirectional Mendelian randomization analyses provided preliminary evidence in support of a causal relationship between (reduced) thalamic NDI and (increased) risk of schizophrenia.

**Conclusions:** Risk-related reduction in NDI is plausibly indicative of reduced density of myelinated axons and dendritic arborization in large-scale cortico-subcortical networks. Cortical, subcortical and white matter micro-structure may be linked to the genetic mechanisms of schizophrenia.

## Introduction

A substantial genetic contribution to the pathogenesis of schizophrenia is indicated by twin and familial heritability estimates of approximately 80% [1, 2, 3], and SNP heritability of approximately 24% [4]. The most recent genome-wide association study (GWAS) identified 270 risk-associated loci, allowing the construction of polygenic risk scores (PRS) that explain up to 7.7% of the variance in schizophrenia [4]. Polygenic risk scores are normally distributed in the general population and have been used to investigate the shared genetics between schizophrenia, neurodevelopmental trajectories and brain morphology [5, 6].

The genetic liability for schizophrenia is thought to cause proximal changes in brain structure and function [7], which then result in distal changes in psychological function and clinical symptoms characteristic of schizophrenia [8, 9, 10, 11]. Although brain structural abnormalities have been consistently reported in schizophrenia case-control studies [12, 13, 14], and are substantially heritable [15, 16, 17], the current evidence linking magnetic resonance imaging (MRI) markers of brain structure to polygenic risk for schizophrenia is inconsistent [18]. Recent studies reported no significant associations between PRS and volume of subcortical nuclei [19, 20, 21], or between PRS and white matter structure [19]. Some studies have reported significant negative associations between PRS and global brain measures, such as total white matter volume (WMV) [22]) or mean cortical thickness (CT) [6]; but effect sizes have been small (*R*^2^ = 0.2%) and not consistently significant between studies [18, 19]. Thickness of insular cortex was specifically associated with polygenic risk for schizophrenia (*R*^2^ = 0.2%) [6] but most prior studies have not found significant associations between PRS and regional brain anatomy [18].

The current lack of clear evidence for an association between genetic risk for schizophrenia and brain structure could be attributable to the relatively small sample sizes of prior genetic neuroimaging studies (100 < N < 15,000), which were likely under-powered to detect small polygenic effects [6, 18, 19, 21]. Inherently low power is necessarily exacerbated when type 1 error is appropriately adjusted to control for multiple testing of regional phenotypes [6]. It is also notable that cortical and subcortical MRI phenotypes so far investigated have mostly been macro-structural metrics, e.g., cortical thickness, which are coarse-grained compared to the predicted effects of risk genes on tissue composition and cellular organization.

Case-control studies of schizophrenia have consistently reported significant reductions in regional cortical thickness (CT), surface area (SA) [13], grey matter volume (Vol)[23], intrinsic curvature (IC, a metric of local connectivity of cortex) [24] and local gyrification index (LGI, a metric of cortical folding) [25]. These are all macro-structural markers of brain morphology that are derived from T1-weighted MRI data and estimated each cortical or subcortical region. In contrast, micro-structural MRI phenotypes are derived from diffusion-weighted imaging (DWI) data, providing finer-grained information about the cellular composition of brain tissue [26, 27, 28]. Case-control studies of schizophrenia have increasingly reported significant differences in grey matter micro-structure (26), such as increased mean diffusivity (MD) [28, 29, 30], decreased fractional anisotropy (FA) [31], decreased neurite density index (NDI) [26] and orientation dispersion index (ODI) [32]. Significant micro-structural abnormalities of white matter tracts have also been reported, including decreased FA [14, 33] and NDI [34, 35], and increased MD [14], in schizophrenia case-control studies.

We therefore hypothesized that macro- and micro-structural MRI markers of grey and white matter would be associated with polygenic risk for schizophrenia. We analysed multimodal MRI and genotype data from N ∼ 30,000 participants in the UK Biobank to conduct a comprehensive study of PRS association with nine brain MRI phenotypes (**Fig.1**). In grey matter, we measured five macro-structural (CT, SA, Vol, LGI and IC) and four micro-structural (FA, MD, NDI and ODI) MRI metrics in 180 bilateral cortical areas; and a subset of these metrics (Vol, FA, MD, NDI and ODI) in 7 subcortical regions. In white matter, we measured FA, MD, NDI and ODI in 15 major axonal tracts. We used the largest available GWAS dataset for schizophrenia (N = 69,369 cases and N = 236,642 controls) to construct schizophrenia PRS for each subject [4]. We expected that this combination of increased sample size for PRS estimation, and MRI measurement of both macro- and micro-structural metrics, would enhance statistical power to test the hypothesis that MRI phenotypes are associated with schizophrenia PRS in the population. In light of the significant associations we discovered by these analyses, we were stimulated to conduct post-hoc analyses of the causal relationships between a subset of NDI metrics and schizophrenia, using Mendelian randomization and genetic correlations.

**Figure 1.**
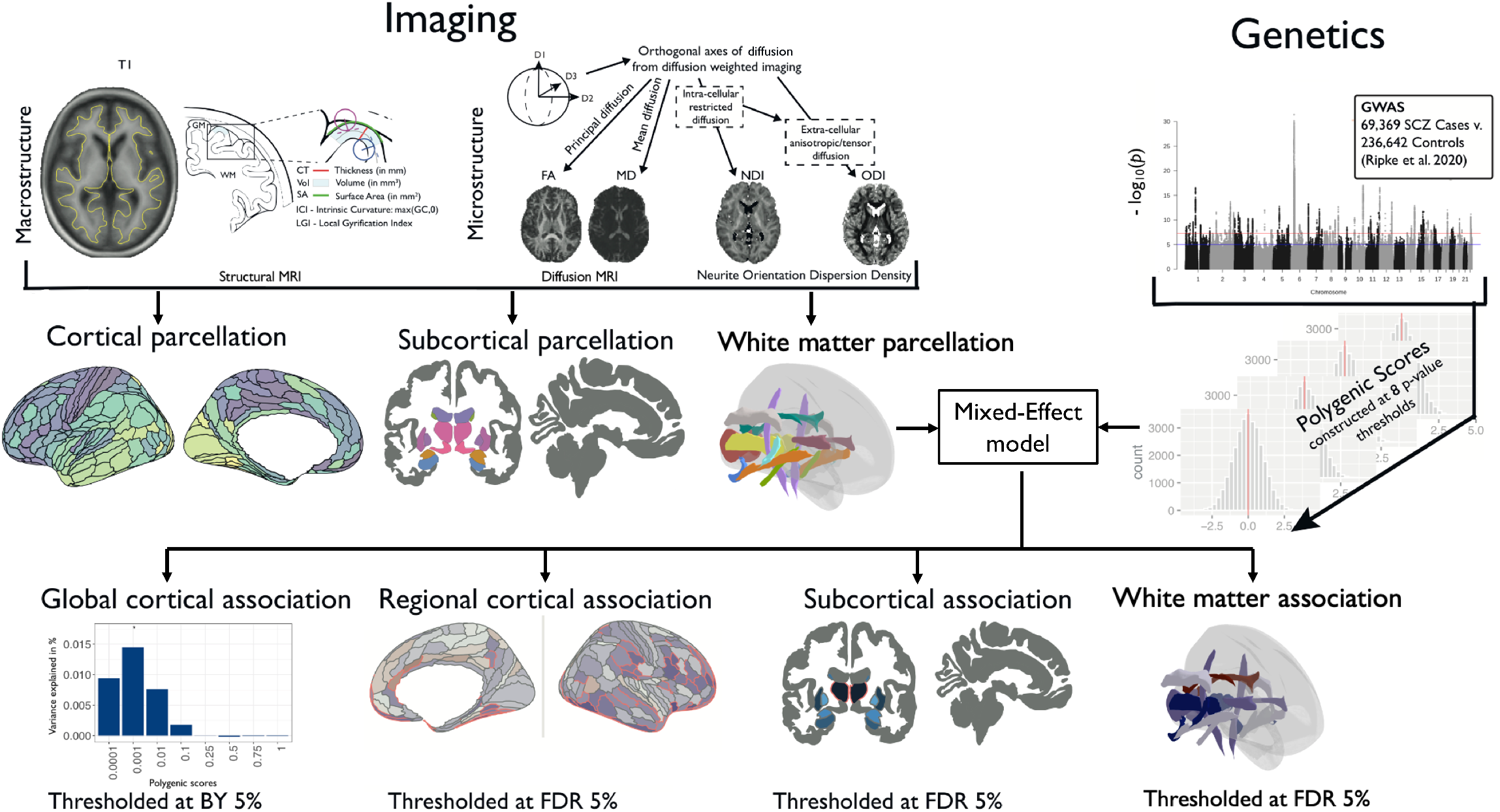
Schematic summary of the study. We estimated five macro-structural metrics and four micro-structural metrics at each of 180 bilateral cortical areas, and on average over all cortical areas, for variable numbers of subjects in the UK Biobank for whom quality-controlled data were available: for cortical thickness (CT), N = 29, 778; surface area (SA), N = 29, 777; grey matter volume (Vol), N = 29, 778; intrinsic curvature (IC), N = 29, 676; local gyrification index (LGI), N = 27, 086; fractional anisotropy (FA), N = 28, 232; mean diffusivity (MD), N = 28, 165; neurite density index (NDI), N = 27, 632; and orientation dispersion index (ODI), N = 27, 658. We also estimated one macro-structural metric and four micro-structural metrics at each of seven subcortical regions (amygdala, accumbens, caudate, putamen, pallidum, hippocampus, and thalamus): Vol, N = 29, 854 − 29, 878; FA, N = 28, 192 − 28, 238; MD, N = 27, 664 − 28, 154; NDI, N = 27, 590 − 27, 638; and ODI, N = 27, 600 − 27, 658. Additionally, we measured four micro-structural metrics at 15 major white matter tracts: FA, N = 27, 987−28, 346; MD, N = 28, 327−28, 346; NDI, N = 28, 247−28, 337; ODI, N = 28, 219−28, 346. Polygenic risk scores for schizophrenia (PRS) were based on GWAS data from 69, 396 cases and 236, 642 controls and were calculated for each participant at eight *P*_*SNP*_-value thresholds for inclusion of significant variants, using the clumping and thresholding approach [36]. To assess associations between multiple schizophrenia polygenic risk scores and cortical phenotypes, we first used mixed effect models to identify which *P*_*SNP*_-value threshold(s) produced the PRS most strongly associated with each cortical metric at global scale (Benjamini-Yekutieli corrected 5%). We then used mixed effect models to test the association between the globally most predictive PRS for each metric at each cortical area, controlling for multiple comparisons with the false discovery rate (FDR) at 5%. For subcortical structures and white matter tracts, we tested for association between each MRI metric and each of eight polygenic risk scores, with FDR = 5%. The Manhattan plot is for illustrative purposes only and based on a previously published schizophrenia GWAS [37], downloaded from url: https://www.med.unc.edu/pgc/pgc-workgroups/.

## Methods and Materials

### Participants

Data were provided by the UK Biobank, a population-based cohort of >500,000 subjects aged between 39 and 73 years [38]. We focused on a subset of N = 40,680 participants for each of whom complete genotype and multimodal MRI data were available for download (February 2020); **Fig.S2**. We excluded participants with incomplete MRI data, or with a diagnosis of schizophrenia (self-reported or by ICD-10 criteria). Prior to analysis of each MRI phenotype, we additionally excluded participants who were robustly defined as outliers by global or regional metrics more than 5 times the median absolute deviation from the sample median (± 5 MAD), see **SI Methods Sample Selection**. Ethical approval was obtained from the Human Biology Research Ethics Committee, University of Cambridge (Cambridge, UK).

### Imaging acquisition and preprocessing

MRI data acquisition has been described in detail elsewhere [39]. Minimally processed T1- and T2-FLAIR-weighted MRI data (and DWI data) were downloaded from UK Biobank (application 20904) ^1^ and further processed with Freesurfer (v6.0.1) [40] using the T2-FLAIR weighted images to improve pial surface reconstruction. Pre-processing steps included bias field correction, registration to stereotaxic space, intensity normalization, skull-stripping, and grey/white matter segmentation; Following reconstruction, the Human Connectome Project (HCP) parcellation [41] was aligned to each individual image and regional metrics were estimated for 180 bilateral cortical areas and seven bilateral subcortical structures. DWI data were co-registered with the T1-aligned parcellation template to estimate FA and MD at each region. Neurite orientation dispersion and density imaging (NODDI) reconstruction was performed using the AMICO pipeline [42]. Probabilistic tractography was used to estimate FA, MD, NDI and ODI values at each of 15 major white matter tracts defined using AutoPtx [43]. Documentation and code for these processing pipelines is available on Github ^2^.

### Genotyping and genetic quality control

Genome-wide genotype data were available for N = 488,377 participants, with DNA acquisition, imputation and quality control pipelines as described elsewhere [44]. We excluded participants if they were not primarily of white British ancestry based on genetic ethnic grouping and subjects with excessive genetic heterozygosity, genotyping rate ≤ 95%, mismatch between reported and genetic sex, and genetic relatedness *>* 0.25 between participants [45]. We also excluded single nucleotide polymorphisms (SNPs) with minor allele frequency ≤ 0.01, that were not in Hardy-Weinberg equilibrium (p ≤ 1 ×10^−6^), variant call rate ≤ 98%, and an imputation quality score ≤ 0.4, resulting in 5,366,036 SNPs.

### Polygenic risk scores

A PRS is calculated by multiplying the number of risk alleles by the effect size of each allele and summing the products over all SNPs for each individual [46]. We used the computationally efficient P-value clumping and thresholding method in PRSice 2 [4, 36, 46, 47]. First, SNPs were clumped using the UK Biobank data as a reference panel so that only the most strongly associated SNP in a region was retained (*r*^2^ = 0.1, physical distance = 250 kb) [36]. Second, for each participant, we estimated a set of eight PRS with varying SNP-wise probability thresholds for inclusion (0.0001 ≤*P*_*SNP*_ ≤ 1) to balance signal to noise ratio [6, 48, 49].

We estimated PRS’s for N = 29,879 participants, using effect sizes for each allele from a trans-ancestry GWAS with N = 69,369 schizophrenia cases and N = 236,642 healthy controls, mostly (∼80%) of European ancestry [4]. Previous studies that used UK Biobank data to test association of MRI markers with polygenic risk estimated PRS from smaller GWAS datasets of both European and East Asian ancestry [4, 6, 19]. Thus, we estimated eight PRS from the full trans-ancestry GWAS [4] for each participant and found that scores were normally distributed at all *P*_*SNP*_ inclusion thresholds (**Figure S3, SI Methods**); the number of SNPs included in the PRS calculation at each probability threshold is reported in **Table S1**.

### Statistical analysis

Statistical analyses were conducted in R [50]. We used linear mixed effect (LME, “nlme” package version 3.1-144) models (Equation 1) to estimate the associations between the scaled PRS and each of the scaled MRI phenotypes with covariates including age, *age*^2^, sex, genotype batch, 15 genetic principal components (Data-Field 22006), and x, y and z coordinates of head position in the scanner to control for static-field heterogeneity (Data-Field 25756-25758) [51]. Hemisphere was fitted as a random effect, resulting in 180 bilateral regions [6, 19, 52], after testing for PRS-by-hemisphere interactions had demonstrated that none were significant at FDR = 5% (**Tables S2-S4**).

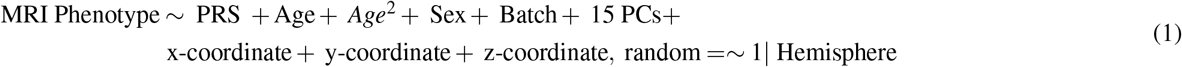

To identify the optimal *P*_*SNP*_ thresholds for PRS construction, and to minimize the multiple testing required for comprehensive brain regional mapping of genetic associations, we decided a priori on a two-step analysis for cortical phenotypes. First, we estimated and tested the association of all eight polygenic risk scores with each MRI metric at a global cortical scale, using the Benjamini-Yekutieli procedure (BY = 5%) to control for multiple tests over all metrics and thresholds. We thus identified the genome-wide probability threshold for PRS inclusion, *P*_*SNP*_-global, that produced the polygenic risk score most strongly associated with each global cortical metric. Second, we used the PRS defined by *P*_*SNP*_-global to test for genetic association with the corresponding metric at a regional scale in 180 cortical areas, covarying for total intracranial volume, with false-discovery rate (FDR) = 5%.

For analysis of the smaller number of subcortical regions (7 regions), and white matter tracts (15 tracts), we tested for association with all eight PRSs at each region, with FDR = 5%, to control for the multiple tests entailed (56 and 120, respectively).

We performed various sensitivity analyses: First, we repeated the regional cortical analysis using the PRS scores at all other *P*_*SNP*_ thresholds and showed that significant associations between PRS and regional MRI phenotypes were largely conserved across *P*_*SNP*_ thresholds (**Figure S4-S12**). Second, we covaried for the corresponding global metric in the LME model of PRS associations with regional metrics, which revealed a highly correlated pattern for regional cortical associations (**Figure S13**). Third we repeated cortical analysis using PRS scores from GWAS data in European samples and showed that our findings were not biased by population stratification (**Figure S14-16**).

We report the proportion of explained phenotypic variance by the PRS in percentages (*R*^2^) and standardized effect sizes (*β*) throughout.

### Bidirectional Mendelian randomization analyses and genetic correlations

We conducted exploratory bidirectional Mendelian randomization (MR) analyses for a subset of imaging phenotypes using the ‘twosampleMR’ package v0.5.6 in the UK Biobank sample [53]. Mendelian randomization employs genetic instruments to test whether there is a causal effect of the exposure phenotype on the outcome phenotype. We tested two directions of causal relationship: (i) schizophrenia (exposure) causing brain NDI changes (outcome); and (ii) brain NDI changes (exposure) causing schizophrenia (outcome). We restricted MR analysis to a subset of global and regional NDI metrics that were most strongly and robustly associated with schizophrenia PRS and showed ≥ 10 genome-wide significant loci, to ensure reasonable statistical power (**Tables S5-S6**).

For MR analysis (i) of the causal effect of schizophrenia on NDI, we used the same GWAS summary statistics as for the PRS analyses [4]. Genetic instruments were chosen at a P threshold of 5 × 10^−8^ and clumped with a distance of 10,000 kb and LD r2 of 0.001. These SNPs were then identified within the outcome GWAS and SNP effect of exposure and outcome data were harmonised to match the effect alleles leading to 184 genetic instruments and 41 imaging phenotypes (**Table S7**).

For MR analysis (ii) of the causal effect of NDI on schizophrenia, we performed GWAS using fastGWA with sample sizes in the range N = 31,722 - 31,760 for different regional metrics (**SI Methods**) [54]. Genetic instruments were chosen at two significance thresholds. Firstly, we used a genome-wide significant threshold of P = 5 × 10^−8^ and clumped with a distance of 10,000 kb and LD r2 of 0.001. Based on these parameters, 40 NDI phenotypes were included and the number of genome-wide significant loci ranged from three to 29 after harmonising the data (**Table S8**). Secondly, we identified genetic instruments for the same 40 NDI phenotypes with a lower GWAS significance threshold of P = 5 × 10^−6^, as the smallest number of genetic instruments after data harmonising was three [48, 55] (**Tables S5-S6**). This increased the number of genetic instruments, which ranged from 21 to 71 after data harmonising (**Table S9**), which was expected to enhance statistical power for MR analysis at the cost of somewhat less stringent type 1 error control in construction of the instrumental variables.

To fit the models, we used inverse variance-weighted Mendelian randomization (IVW), which assumes that all SNPs are valid genetic instruments, as the main method to estimate causal effects [56]. The robustness of significant findings obtained using IVW was assessed using two additional methods (sensitivity analyses): weighted median method (WM), which provides consistent results even when 50% of the genetic instruments are invalid; and MR-Egger, which accounts for pleiotropy [56], and we tested for heterogeneity and horizontal pleiotropy.

To evaluate consistency in effect direction, we additionally calculated genetic correlations between schizophrenia and the same 41 imaging phenotypes using Linkage Disequilibrium Score Regression v1.0.1 [57] and corrected for multiple comparison using FDR.

## Results

### Sample

The sample sizes available after QC for analysis of cortical, subcortical and white matter structure varied between regions and MRI metrics in the range N = 27,086-29,878 (**Fig.1**). All samples comprised approximately 55% female, 45% male participants aged 40-70 years, with mean age ∼ 55 years; see **Tables S10 – S12** for details.

### Global cortical MRI phenotypes

Two global cortical metrics were significantly associated with PRS at one or more *P*_*SNP*_-value thresholds (BY = 5%). Both were micro-structural phenotypes derived from DWI or NODDI: FA was significantly negatively associated with the PRS constructed with *P*_*SNP*_ ≤ 0.001; and NDI was significantly negatively associated with PRS constructed at *P*_*SNP*_’s ≤ 0.0001, 0.001 and 0.01 (**Fig.2 A, Table S13**). These results indicate that participants with increased polygenic risk for schizophrenia have decreased global fractional anisotropy and neurite density index “on average” over the whole cortex. The association between PRS and NDI was more robust to the choice of probability threshold used to construct the risk score; and PRS accounted for a larger proportion of the variance in NDI (∼0.05%), compared to FA (∼0.02%) and other metrics.

**Figure 2.**
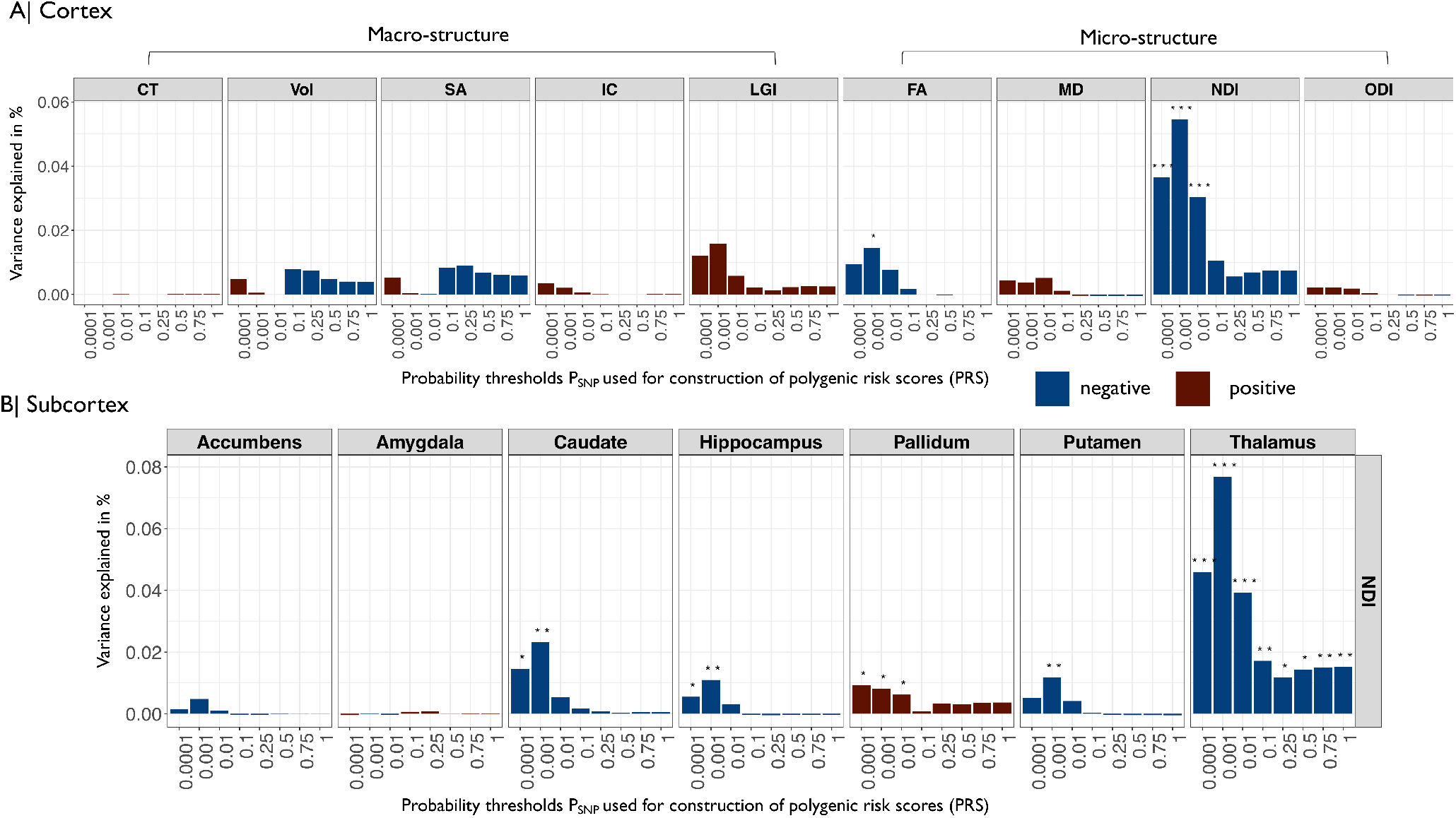
Associations between polygenic risk scores for schizophrenia and global cortical (A) and regional subcortical (B) metrics of human brain structure. (A) Barcharts of variance explained by schizophrenia PRS (*R*^2^, *y*-axis) constructed at each of eight probability thresholds (0.0001 ≥ *P*_*SNP*_ ≤ 1, *x*-axis) for each of nine global mean cortical metrics: CT, cortical thickness; GMV, grey matter volume; SA surface area; IC intrinsic curvature; LGI local gyrification index; FA fractional anisotropy; MD mean diffusivity; NDI neurite density index; ODI orientation dispersion index. Blue bars indicate negative associations and red bars positive associations; asterisks indicate P-values for association after FDR correction: (∗*P*≤ 0.05, ∗∗*P*≤ 0.01, ∗∗∗ *P* ≤ 0.001). Polygenic risk scores for schizophrenia were significantly negatively associated with global neurite density index and fractional anisotropy. (B) Barcharts of variance explained by PRS (*R*^2^, *y*-axis) constructed at each of eight probability thresholds (0.0001 ≥ *P*_*SNP*_ ≤ 1, *x*-axis) for NDI measured at each of seven subcortical regions (colours and asterisks code sign and significance of association as in A). PRS was significantly negatively associated with NDI in thalamus, hippocampus, putamen and caudate; and significantly positively associated with NDI in pallidum.

### Regional cortical MRI phenotypes

The macro-structural metrics (CT, SA, Vol) were positively correlated with each other and negatively correlated with LGI. Among the micro-structural metrics, NDI and FA were positively correlated; and both were negatively correlated with MD and ODI (**Fig.3A**).

**Figure 3.**
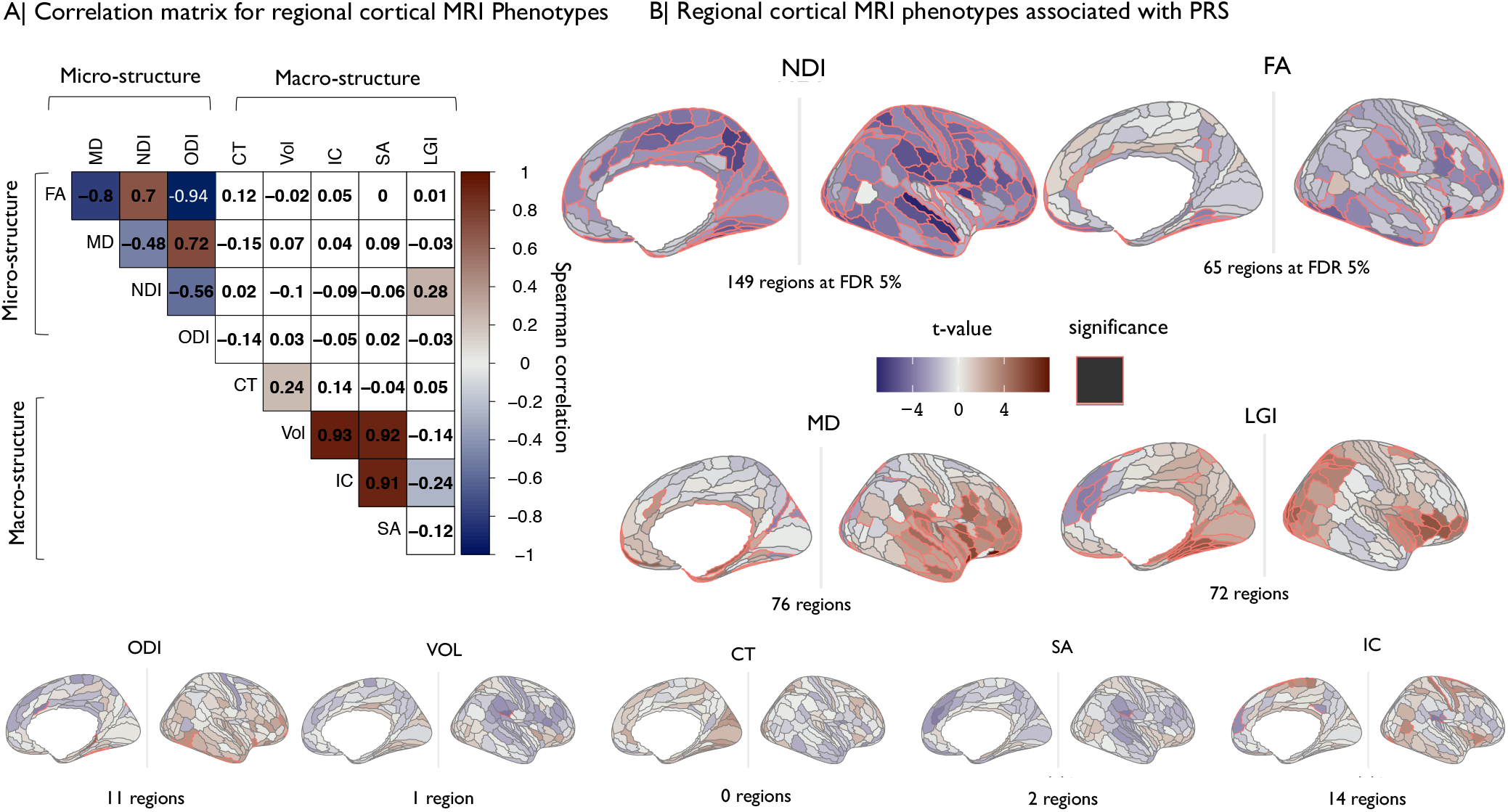
Regional cortical MRI phenotypes: correlations between metrics and associations of each metric with schizophrenia polygenic risk scores. (A) Matrix of Spearman’s correlation for each pair of nine MRI metrics. Shades of blue indicate significant negative correlation and shades of red indicate significant positive correlations. (B) Cortical *t*-maps representing strength of association between schizophrenia PRS and regional MRI phenotypes; regions where the effect of PRS is statistically significant at FDR = 5% are outlined in red. NDI and FA metrics, which were globally decreased by genetic risk for schizophrenia (**Fig.2**), were significantly regionally decreased in multiple areas. MD and LGI metrics were significantly regionally increased by genetic risk in several areas.

There were significant associations between PRS and eight out of nine MRI metrics in at least one cortical area, with the exception being CT (**Fig.3B**). The proportion of regional variance explained by PRS varied between metrics in the range ≤ .002% *R*^2^ ≤ .08% with -.03 ≤ *β* ≤ +.03; see **Table S14**.

Among the micro-structural metrics, NDI was significantly associated with PRS in 149 cortical areas (maximum *R*^2^ = .06%, *β* = -0.03). The top ten areas where NDI was most strongly negatively associated with PRS were located in association auditory cortex, early auditory cortex, ventral visual stream, posterior cingulate cortex, insular and frontal opercular cortex, posterior opercular cortex, inferior parietal cortex and superior parietal cortex (see **Table S15** for details). FA was negatively associated with PRS in 63 regions (maximum *R*^2^ = .01%, *β* = -0.01); and positively associated with PRS in two regions (maximum *R*^2^ = .01%, *β* = .01). The top ten regions where FA was most strongly associated with PRS were all areas of temporal, cingulate, frontal and insular cortex, where decreased FA was associated with higher risk scores. MD was significantly associated with PRS in 76 regions (66 positive and 10 negative associations). ODI was the micro-structural metric least frequently associated with PRS, at 11 cortical regions.

Of the macro-structural metrics, LGI showed the highest number of significant associations with PRS (72 regions; 67 positive and five negative) compared to Vol, SA and IC, each of which was significant in less than 15 regions (maximum *R*^2^ ≤ .02%, maximum |*β* | ≤ .01).

Since the MRI phenotypes were not independent of each other (**Fig.3 A**), we further explored the spatial co-localisation of 187 genetic associations with different cortical metrics. The cortical t-maps of PRS association with NDI and FA were significantly 188 positively correlated with each other, and were negatively correlated with the cortical t-maps of PRS associations with ODI 189 (both NDI and FA) and MD (FA only) (**Fig.4A**). In other words, regions where genetic risk was most strongly associated with decreased FA (FA(*t*) ≪ 0) tended to be the same regions where PRS was most strongly associated with decreased NDI (NDI(*t*) ≪ 0) and increased MD (MD(*t*) ≫ 0) (**Fig.4** B).

**Figure 4.**
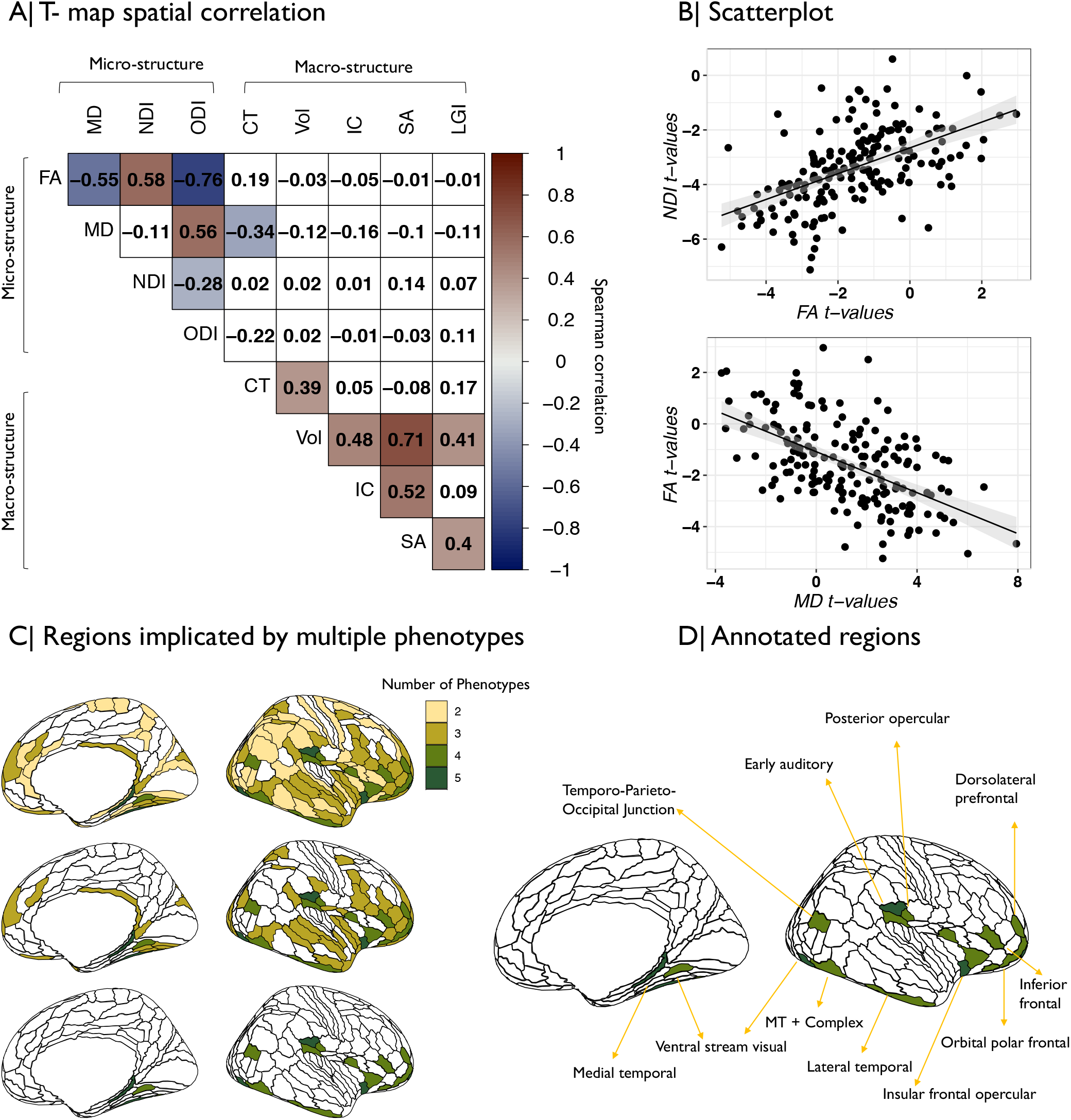
Anatomical co-localisation of polygenic risk effects for schizophrenia on multiple MRI phenotypes. (A) Matrix of spatial correlations between cortical *t*-maps of association between schizophrenia PRS and nine MRI metrics. (B) Scatterplot showing the relationships between (top) cortical *t*-maps of FA and NDI and (bottom) cortical *t*-maps of FA and MD; each point represents a cortical area. (C) Cortical map colour coded to indicate the number of MRI phenotypes that were significantly associated with schizophrenia PRS at each region. Coloured regions had at least two (top), three (middle), or four (bottom), MRI phenotypes significantly associated with genetic risk of schizophrenia. (D) The brain regions where schizophrenia PRS was significantly associated with four MRI phenotypes were anatomically located in medial and lateral temporal cortex, ventral visual stream, insular and frontal cortex.

We also simply counted the number of different MRI phenotypes that were significantly associated with PRS at each of 180 cortical areas. There were genetic effects on at least two metrics in 122 regions (**Table S16**), at least three metrics in 68 regions, and at least four metrics in 21 regions (**Fig.4** C). The most frequently co-localised genetic associations were with NDI and MD (63 regions) and NDI and FA (61 regions) (see **SI Results**). Regions that were associated with PRS in terms of multiple MRI phenotypes were located in (inferior) frontal, insular, temporal, auditory and ventral visual stream areas of cortex (**Fig.4D**, **Table S16**).

### Regional subcortical MRI phenotypes

We estimated the association between PRS and one macro-structural (Vol) and four micro-structural phenotypes (MD, FA, NDI, ODI) at seven subcortical regions. The single strongest effect was a negative association between PRS and NDI of thalamus (*R*^2^ = .08%, *β* -.03). NDI was also negatively associated with PRS in the caudate, hippocampus, and putamen; and positively associated with PRS in pallidum (**Fig.2B, Table S17**). Further significant associations were found between PRS and MD of the amygdala, caudate, hippocampus, pallidum and putamen; between PRS and Vol of caudate, putamen and hippocampus; between PRS and FA of putamen and thalamus; and between PRS and ODI of pallidum and thalamus **(Figure S17)**.

### Regional white matter phenotypes

We identified a large number of significant associations between PRS and micro-structural metrics in 15 white matter tracts: NDI in 14 out of 15 tracts, FA in 12 tracts, MD in 11 tracts, and ODI in 5 tracts (**Fig.5, Fig. S19**). These results indicate that higher genetic risk for schizophrenia is associated with decreased NDI and FA, and increased MD, within callosal fibers, projection fibers, association fibers, limbic system fibers and brainstem tracts. The highest proportion of phenotypic variance explained by PRS was found for NDI of the forceps minor (*R*^2^ = .12%, *β* = -.04) followed by NDI of projection fibers (superior thalamic radiation *R*^2^ = .11%, *β* = -.03) and association fibers (inferior fronto-occipital fasciculus *R*^2^ = .10%, *β* = -.03). Those associations showed a consistent direction of effect and were significant at all *P*_*SNP*_ inclusion thresholds. The strongest negative association between PRS and FA was within the forceps minor (*R*^2^ = .11%, *β* = -.03), followed by projection fibers (superior thalamic radiation *R*^2^ = .06%, *β* = -.02), and association fibers (anterior thalamic radiation *R*^2^ = .06%, *β* = -.02). The strongest positive associations between PRS and MD were located in association fibers (uncinate fasciculus *R*^2^ = .07%, *β* = .03) followed by limbic system fibers (cingulate gyrus part of cingulum R2 = .04%, *β* = .02) and the forceps minor (*R*^2^ = .03%, *β* = .02) (**Table S18**).

**Figure 5.**
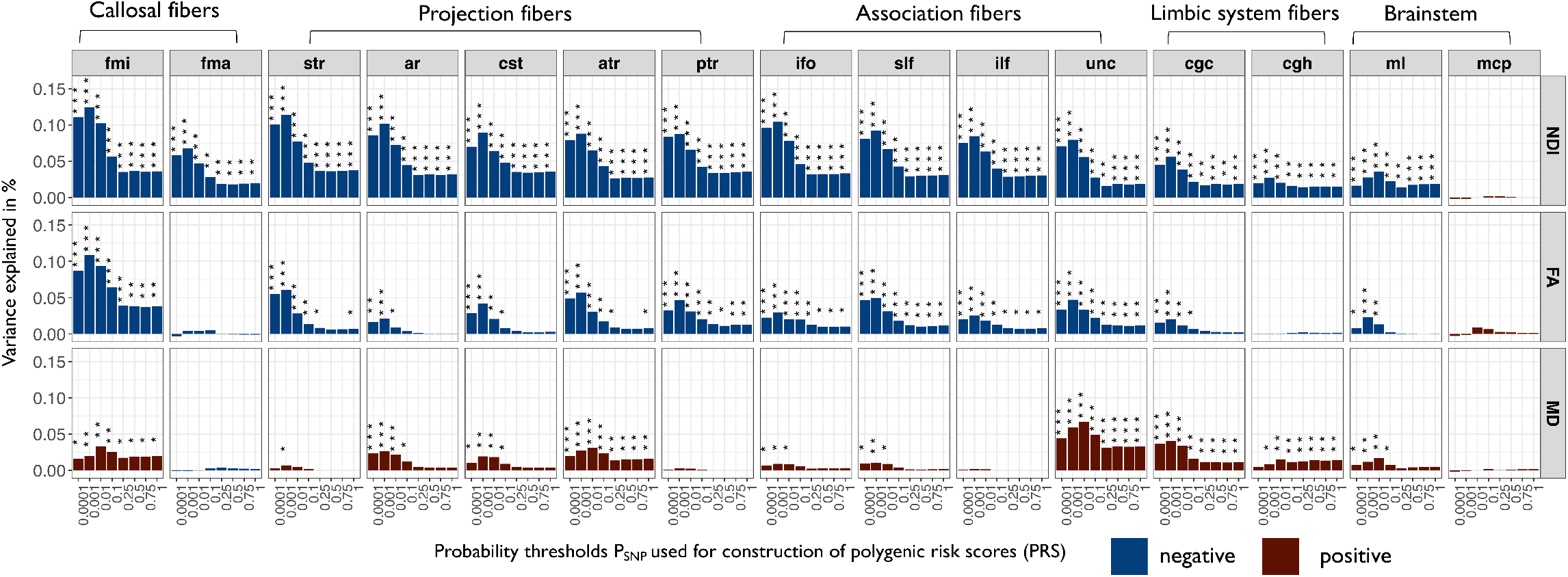
Associations between polygenic risk scores for schizophrenia and white matter tracts. Barcharts of variance explained by schizophrenia PRS (*R*^2^, y-axis) constructed at each of eight probability thresholds (0.0001 ≥ *P*_*SNP*_ ≤ 1, x-axis) for each of three white matter metrics (NDI neurite density index; FA fractional anisotropy; MD mean diffusivity) measured at 15 major white matter tracts: mcp, middle cerebellar peduncle; ml, medial lemniscus; cst, corticospinal tract; ar, acoustic radiation; atr, anterior thalamic radiation; str, superior thalamic radiation; pts, posterior thalamic radiation; slf, superior longitudinal fasciculus; ilf, inferior longitudinal fasciculus; ifo, inferior fronto-occipital fasciculus; unc, uncinate fasciculus; cgc, cingulate gyrus part of cingulum; cgh, parahippocampal part of cingulum; fmi, forceps minor; and fma, forceps major. Blue bars indicate negative associations and red bars positive associations; asterisks indicate P-values for association after FDR correction: * P 0.05, ** P ≤ 0.01, *** P ≤ 0.001. Polygenic risk scores for schizophrenia were significantly associated with NDI, FA and MD of multiple white matter tracts.

### Bidirectional Mendelian randomization on schizophrenia and NDI metrics

For these post-hoc analyses of causality, we focused on 41 NDI phenotypes (global NDI and regional NDI metrics in 21 cortical areas, 5 subcortical nuclei, and 14 white matter tracts). After correction for multiple comparisons, there was no significant evidence for a causal effect of schizophrenia on NDI metrics (MR (i)), and there was no significant evidence for a causal effect of NDI metrics on schizophrenia (MR (ii)) using genetic instruments identified by genome-wide association significance threshold of P = 5 × 10^−8^ (**Tables S7, S8**). However, for MR (ii), when we relaxed the inclusion threshold for genetic instruments to P = 5 × 10^−6^, we found that lower genetically predicted NDI in the thalamus was associated with increased genetically predicted risk for schizophrenia (IVW method *β* = -0.16, *P*_*FDR*_ = 0.02), suggesting that reduced thalamic NDI may cause increased risk of schizophrenia. Sensitivity analyses using the weighted median method were also significant (*β* = -0.14, P = 0.002). NDI of thalamus showed significant heterogeneity between individual genetic variants (P for Q-test < 0.0001) but the Egger intercept was not significant, suggesting no evidence for substantial pleiotropic effects (P = 0.42) (**Table S9**). Additionally, leave-one-out analysis did not indicate that the result was driven by any one genetic variant (**Figure S20**).

Genetic correlations were modest and did not survive multiple testing correction. However, 80% of the correlations were in the negative direction predicted by significant negative associations between NDI and PRS scores (**Table S19**).

## Discussion

We estimated the strength of association between polygenic risk scores for schizophrenia and nine MRI phenotypes measured at 180 cortical areas, seven subcortical structures and 15 white matter tracts. In strong support of our motivating hypothesis, we found that PRS was significantly associated with micro-structural metrics of brain tissue composition at global and regional scales of cortex; in the thalamus, basal ganglia and hippocampus; and extensively in white matter tracts. First we interpret and integrate these signals of significant association between genetic risk for schizophrenia and brain MRI metrics, especially NDI; then we discuss post-hoc analyses of causality stimulated by these results.

### Neurite density index - a plausible brain MRI marker for schizophrenia risk?

Of all nine MRI metrics considered, NDI was the most robustly associated with PRS. NDI is derived from NODDI, an MRI sequence that was developed to estimate the micro-structural complexity of dendrites and axons - collectively referred to as neurites - in grey and white matter of the living brain. NODDI compartmentalizes tissue into three microstructural environments - intra-cellular, extra-cellular, and cerebro-spinal fluid - each of which has different diffusion properties. The intra-cellular compartment refers to the space bound by neurites and allows for the measurement of their density (NDI) and their spatial orientation (ODI) [58, 59]. *In vivo* estimates of NDI have been biologically validated by *ex vivo*, histological estimates of neurite density in mice [60], and correlated with cortical myelination in humans [59].

Thus, a reasonable interpretation of our results is that polygenic risk for schizophrenia in the general population is associated with decreased axonal and/or dendritic density in grey and white matter. While we cannot ascribe causality using the current methods, this interpretation is consistent with several lines of previous research in individuals with schizophrenia. First, schizophrenia case-control studies have reported abnormally decreased NDI in the cortex, hippocampus and white matter [26, 35]. Secondly, post-mortem studies have reported multiple abnormalities of neurite structure in schizophrenia, including reduced dendritic arborisation [61], reduced spine density [61], reduced axonal myelination [62], and reduced oligodendrocytes [63]. Third, recent GWAS studies have identified risk genes for schizophrenia that are expressed in neurons and implicated in dendritic arborization and microcircuit formation, synaptic plasticity, and glutamatergic neurotransmission [4, 64, 65]. It is intriguing to speculate which individual risk genes might be most relevant to the relationship between PRS and reduced NDI but we cannot certainly resolve this question from these results. Fourth, locally reduced density of myelinated axons and dendrites will likely reflect atypical connectivity between brain structures [62, 66], as anticipated by long-standing theories of schizophrenia as a dysconnectivity syndrome [67, 68, 69, 70].

### Integration of genetic effects across cortical phenotypes

While NDI was the one most strongly associated with PRS, almost all other metrics (except CT) also showed some significant regional associations with genetic risk. We consider that this apparent pleiotropy of genetic effects on brain structure largely reflects correlations between the MRI metrics [27]. For example, NDI was positively correlated with FA, and both NDI and FA were negatively correlated with MD (**Fig. 3**). This is not surprising because increased NDI restricts isotropic diffusion of protons in the tissue water compartment so that FA is increased and MD is decreased [27]. These three metrics are thus complementary measures of the same or similar tissue composition characteristics, which explains the anatomical co-localisation of genetic effects on NDI, FA and MD (**Fig. 4**). These co-localised, multi-metric associations with PRS were concentrated in auditory and lateral temporal cortex, pre-frontal and orbito-frontal cortex, anterior cingulate cortex, and insular cortical areas, many of which have previously been reported to show increased MD and/or decreased FA in case-control studies of schizophrenia [28, 29, 30, 31].

Of the macro-structural phenotypes, LGI was positively correlated with MD and positively associated with genetic risk for schizophrenia. Local gyrification is assumed to capture early neurodevelopmental changes that are relatively stable after birth [25, 71]. Case-control data have recently shown increased LGI in schizophrenia, interpreted as a marker of structural dysconnectivity [25]. It is uncertain whether polygenic effects on macro-structural markers are mechanistically distinct from the effects on NDI and related micro-structural markers. However, micro-structural MRI metrics were clearly more strongly associated with genetic risk for schizophrenia than the macro-structural metrics that have previously been investigated as candidate endophenotypes.

### Genetic risk and subcortical structures

Atypicality in subcortical structures is a frequently reported finding in schizophrenia cases [12], and in their non-psychotic first-degree relatives [72], compared to healthy controls. However, this is the first study to identify significant associations between polygenic risk for schizophrenia and subcortical structures in a population sample. NDI was again the MRI metric most sensitive to genetic association, especially in the thalamus, where PRS was significantly associated with reduced NDI. The thalamus plays a key role in cognitive and emotional processes that are clinically impaired in schizophrenia [73, 74]. Thalamic volume reductions [73] and abnormal functional connectivity between thalamus and cortex [75] have been previously reported in schizophrenia. The basal ganglia (putamen, pallidum and caudate nucleus) and the hippocampus also demonstrated PRS-related changes in NDI and related micro-structural metrics. There was also a robust (negative) association between PRS and hippocampal volume, consistent with extensively replicated case-control differences in hippocampal volume [12].

### Genetic risk and white matter tracts

Abnormalities of white matter micro-structure, consistent with aberrant inter-hemispheric, fronto-temporal and cortico-thalamic connectivity, have been frequently reported in schizophrenia [14, 33]. We identified significant associations between genetic risk for schizophrenia and micro-structural metrics in most of the major axonal tracts we measured, indicating that PRS is associated with widespread disruption or disorganization of white matter. The strongest effect was a negative association between PRS and NDI of the forceps minor, the anterior part of the corpus callosum, inter-hemispherically connecting the frontal lobes. However, PRS was also significantly associated with NDI, FA and MD in intra-hemispheric association fibers, cortico-subcortical projection fibers, limbic system and brainstem fibers, indicating that genetic risk for schizophrenia is associated with anatomically widespread disruption of white matter tracts.

### Investigation of causality

Association between PRS and brain MRI metrics, however statistically significant or anatomically plausible, is uninformative about the underlying direction of causality. In post-hoc analyses restricted to a subset of the NDI metrics most strongly associated with schizophrenia, we used bidirectional Mendelian randomization to test for causal effects of schizophrenia on NDI metrics (MR(i)), and for causal effects of NDI metrics on schizophrenia (MR(ii)). We found no evidence for a significant causal effect of schizophrenia on global grey matter, cortical, subcortical or white matter tract NDI metrics. We also did not find significant causal effects of any NDI metrics on schizophrenia using genetic instruments selected at a stringent GWAS-significant threshold of P = 5 × 10^−8^. However, we did find a significant, potentially causal relationship between lower NDI in the thalamus and increased risk for schizophrenia when we used a less stringent significance threshold to increase the number of genetic instruments and thus boost the power of MR analysis. We consider that this amounts to suggestive evidence that thalamic NDI may causally predict schizophrenia, consistent with the prior observation that thalamic NDI was the regional MRI metric most strongly and robustly associated with PRS for schizophrenia. However, we also consider that the sample size available for GWAS of NDI metrics was under-powered to identify sufficient instrumental variables at the conventional threshold for genome-wide significance (or to estimate genome-wide associations with sufficient efficiency for significant genetic correlation analysis). More definitive investigations of the causal relationship between MRI metrics and schizophrenia, on the basis of better genetic instruments defined by larger MRI samples, will be important to pursue in future [48].

### Strengths and limitations

It is a strength that we used the largest GWAS to date to construct schizophrenia PRS at multiple *P*_*SNP*_-value thresholds for inclusion of risk variants. We also used the largest and most methodologically diverse MRI dataset to date in order to assess PRS associations with brain structure. However, the diagnostic variance in schizophrenia explained by PRS is still relatively small (7.7%) [4] and, in line with previous findings [6, 19], the proportion of variance in cortical and subcortical structures explained by the PRS is even smaller (*<* 1%). It is expected that even larger sample sizes in future studies might implicate other brain structures or MRI metrics than those significantly associated with PRS in this study [4]. The reported results should be more robustly assessed by future translational MRI and histological studies of grey and white matter micro-structure in human post mortem data or animal models of genetic risk for schizophrenia. Finally, the UK Biobank is an ageing cohort of largely European descent that is on average wealthier and healthier than the general population [76]. The generalisabilty of these results should be investigated in more demographically diverse and epidemiologically relevant samples.

### Summary

Polygenic risk scores for schizophrenia were most robustly associated with significant changes in micro-structural MRI metrics in the cortex, the subcortex, and white matter tracts, in a large population sample. These results provide substantial new evidence in support of the pathogenic model that genetic risk for schizophrenia is associated with reduced neurite density, and anatomical dysconnectivity, in cortico-subcortical networks.

## Supporting information

Supplementary Methods & Results

## Data Availability

The present project used imaging and genetic data made available through the UK Biobank database. Derived data will be made available through the UK Biobank access management system. Any code used to generate derived data is shared on GitHub. GWAS summary statistics for schizophrenia can be accessed from the Psychiatric Genomics Consortium .

https://www.med.unc.edu/pgc/

https://github.com/ucam-department-of-psychiatry/UKB

## Acknowledgements

E.-M.S. is supported by a PhD studentship awarded by the Friends of Peterhouse. This research was co-funded by the National Institute of Health Research (NIHR) Cambridge Biomedical Research Centre and a Marmaduke Sheild grant to R.A.I.B. and V.W. E.T.B. is an NIHR Senior Investigator. R.R.G was funded by a Guarantors of Brain Fellowship. R.A.I.B. is supported by a British Academy Post-Doctoral fellowship and the Autism Research Trust. We wish to thank Dr Petra Vertes and Dr Lisa Ronan for their advice on research design and Dr Simon R White for his statistical advice and support. The views expressed are those of the author(s) and not necessarily those of the NHS, the NIHR or the Department of Health and Social Care. This research was possible due to an application to the UK Biobank (project 20904).

## Author contributions statement

E.-M.S., R.A.I.B., V.W., G.K.M. and E.T.B. designed research. J.S. advised on research design. E.-M.S., R.A.I.B., V.W., and R.R.G. analyzed data. E.-M.S. and R.A.I.B. made figures. E.-M.S., R.A.I.B. and V.W. performed research. E.-M.S. and E.T.B. wrote the paper.

## Footnotes

1 https://biobank.ctsu.ox.ac.uk/crystal/crystal/docs/brainmri.pdf

2 https://github.com/ucam-department-of-psychiatry/UKB

