## Supplementary Methods & Results for "Grey and white matter micro-structure is associated with polygenic risk for schizophrenia"

### 1. SI Methods

#### 1.1 Imaging data acquisition

MRI data[1] was collected on a 3T Siemens Skyra scanner (Siemens, Munich, Germany) using a 32-channel receive head coil. T1-weighted images were acquired using a 3D MPRAGE sequence with the following key parameters; voxel size 1x1x1mm, TI/TR = 880/2000 ms, Field-of-view = 208x256x256 matrix, scanning duration: five minutes. The diffusion imaging data was acquired using a monopolar Steejskal-Tanner pulse sequence and multi-shell acquisition (b=0s/mm2, b=1,000s/mm2, b=2,000s/mm2) with the following key parameters; voxel size 2x2x2mm, TE/TR = 92/3600 ms, Field-of-view = 104x104x72 matrix and scanning duration = seven minutes (1).

#### 1.2 Imaging preprocessing

We obtained minimally processed T1 and T2-FLAIR weighted data from the UK Biobank. Minimal processing for T1 weighted data included defacing, cutting down the field-of-view and gradient distortion correction using FSL's Brain Extraction Tool (2) and FLIRT (FMRIB's Linear Image Registration Tool) (3). The data was then nonlinearly warped to MNI152 space using FNIRT (FMRIB's Nonlinear Image Registration Tool) (4). Next, tissue-type segmentation was applied using FAST (FMRIB's Automated Segmentation Tool) (5) and a bias-field-corrected version of the T1 is generated (1). Minimal processing for Diffusion MRI data included correction for eddy currents (6, 7), head motion, outlier-slices removal and gradient distortion correction (1).

#### 1.3 Imaging quality control

We used T1-weighted and T2-weighted scans for the Freesurfer anatomical image reconstruction, this approach has been shown to improve anatomical reconstruction (8). Morphometric measures in subjects without T2 scans were biased, in that average CT was lower compared to subjects

---

[1] https://biobank.ctsu.ox.ac.uk/crystal/crystal/docs/brain_mri.pdf

with both T1 and T2 images **(Fig.S1)**. Thus, we excluded participants without T2 scans from all analyses.

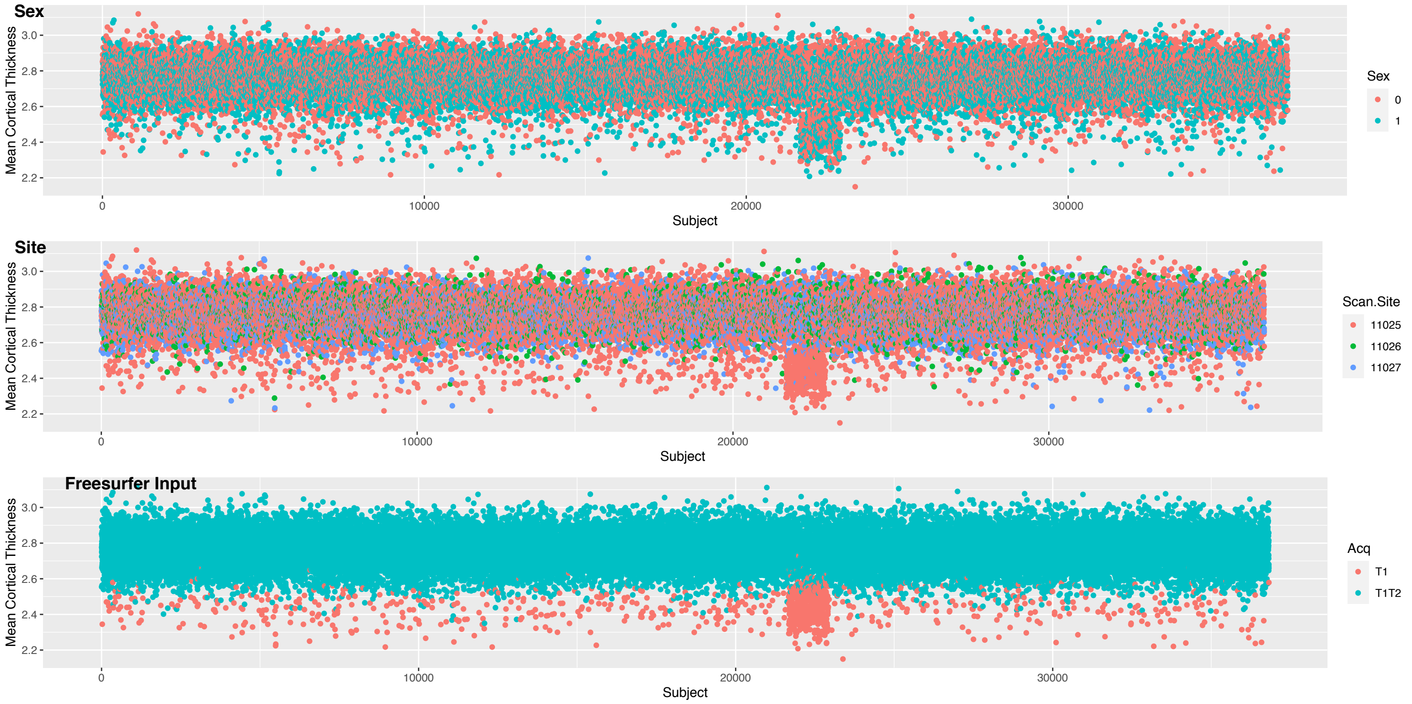

**Figure S1. Quality control.** Basic visual inspection of known sources of noise in Freesurfer reconstruction. Mean cortical thickness (y-axis) is plotted based on sex (male, female), scanning sites and acquisition (availability of T1 or both T1 and T2 scans) for all subjects (x-axis). Mean cortical thickness was biased for subjects that only had T2 scans, which is why we excluded subjects that did not have T2 scans.

##### 1.4 Sample Selection

Participants were recruited by the UK Biobank and included a subset that underwent multi-modal brain imaging (9). At the time of analyses, brain scans of 40,680 participants were available for download. **Fig.S2** clearly outlines how we arrived at the final sample for conducting cortical and subcortical analyses. We excluded subjects with a diagnosis of schizophrenia and subjects that did not survive imaging or genetic quality controls. For cortical analyses we additionally excluded global and regional outliers for each phenotype separately (e.g. a subject that was only an outlier in global CT was excluded for this phenotype but not for other phenotypes). Thus, the final sample size varied between phenotypes with a minimum sample size of 27,086 subjects for LGI and a maximum of 29,778 subjects for CT and Vol. For subcortical analyses we excluded regional outliers for each phenotype and each subcortical structure (e.g. a subject that was only an outlier for Vol of the thalamus was excluded from this sample but not for Vol of the amygdala). For white matter tracts we excluded regional outliers for each phenotype and each tract (e.g. a subject that was an outlier for FA of forceps major was excluded from this sample but not for MD of forceps minor).

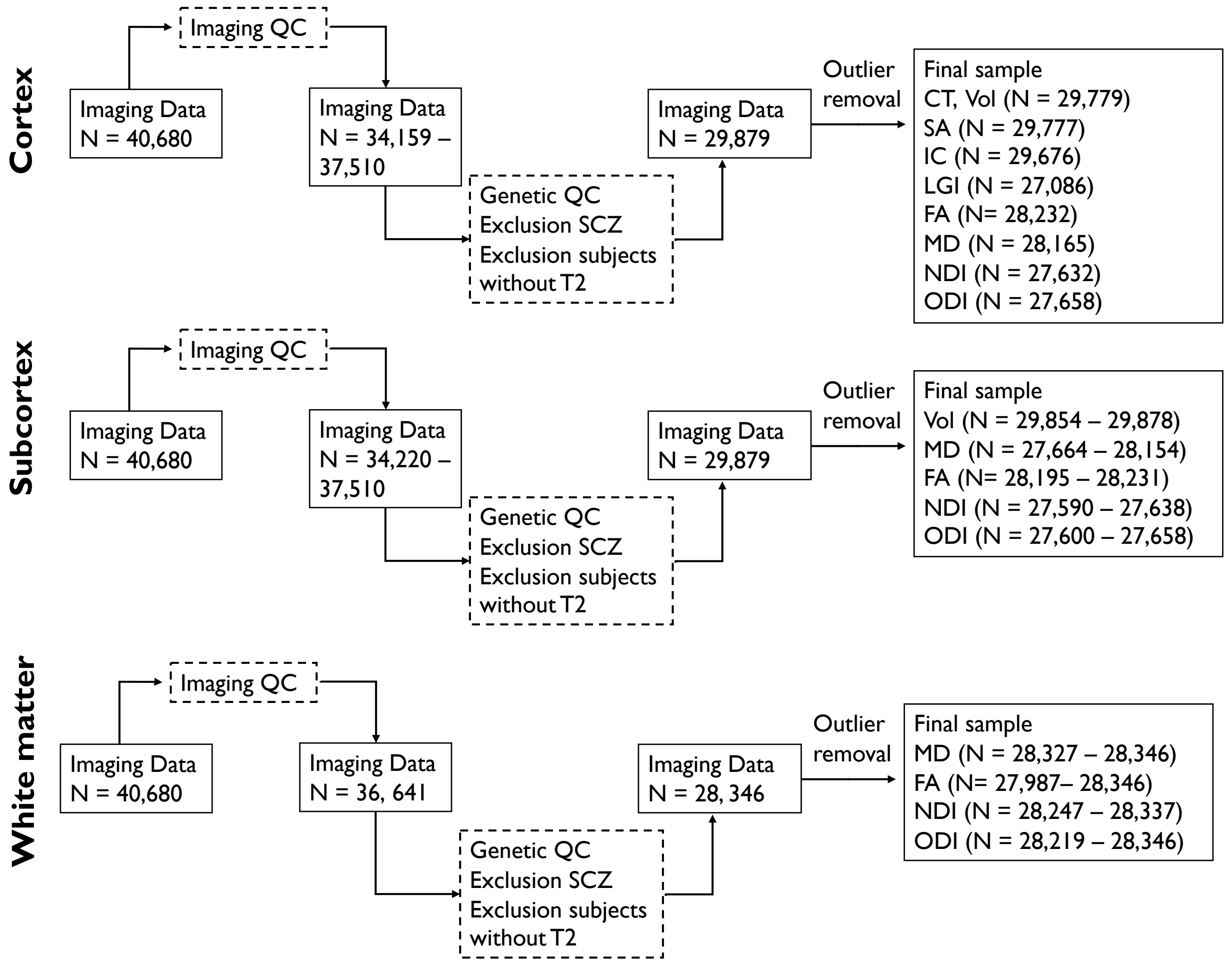

**Figure S2. Sample selection.** This flowchart clearly outlines how we arrived at the final samples for cortical, subcortical and white matter analyses.

##### 1.5 Polygenic risk scores based on European GWAS

As a sensitivity analysis we additionally repeated our analysis using polygenic risk scores based on a GWAS with subjects of European ancestry (10). We followed the analysis pipeline reported in the main paper. The GWAS included 11,308,217 SNPs for 33,640 cases and 43,456 controls. SNPs were clumped so that only the most strongly associated SNP in a region was retained ($R^2$ = 0.1, physical distance = 250 kb). We constructed eight polygenic risk scores with varying $P_{\text{SNP}}$-

value thresholds ($P_{SNP} \leq 0.0001, \leq 0.001, \leq 0.01, \leq 0.1, \leq 0.25, \leq 0.5, \leq 0.75, \leq 1$) for each individual using the clumping and thresholding method in PRSice 2 (11).

##### 1.6 GWAS for neuroimaging phenotypes

All GWAS were conducted using FastGWA (12). FastGWA is a tool for mixed model based GWAS analysis of large-scale data and can simultaneously account for both relatedness and subtle population stratification in the analyses. We included only individuals of self-reported white British ethnicity, and from this group of individuals, excluded individuals who were above +/- 5 SD from the means of the first two genetic principal components, and had a genotyping rate of 95%. Additionally, we excluded individuals whose genetic sex did not match their reported sex and individuals with excessive genetic heterozygosity. We used all genotyped and imputed SNPs in the UK Biobank that had a minor allele frequency > 0.1%, did not deviate from Hardy-Weinberg equilibrium ($P > 1x10^{-6}$), had a genotyping rate of 95%, and, for imputed SNPs, had an imputation $R^2 > 0.4$. All phenotypes were scaled to a mean of 0 and a standard deviation of 1. Prior to analysis of each MRI phenotype, we additionally excluded participants who were robustly defined as outliers by global or regional metrics more than 5 times the median absolute deviation from the sample median (± 5 MAD). For all GWAS, we included age, $age^2$, sex, age x sex, $age^2$ x sex, imaging centre, first 40 genetic principal components, mean framewise displacement, maximum framewise displacement, and Euler Index (13) as covariates. In addition, for structural MRI metrics derived from T1, we included the availability of T2 scans as covariates.

#### 2. SI Results

##### 2.1 Polygenic scores

**Fig. S3** shows the distribution of polygenic risk scores based on the trans-ancestry GWAS at each $P_{SNP}$-value inclusion threshold. Polygenic risks scores were normally distributed within the current sample.

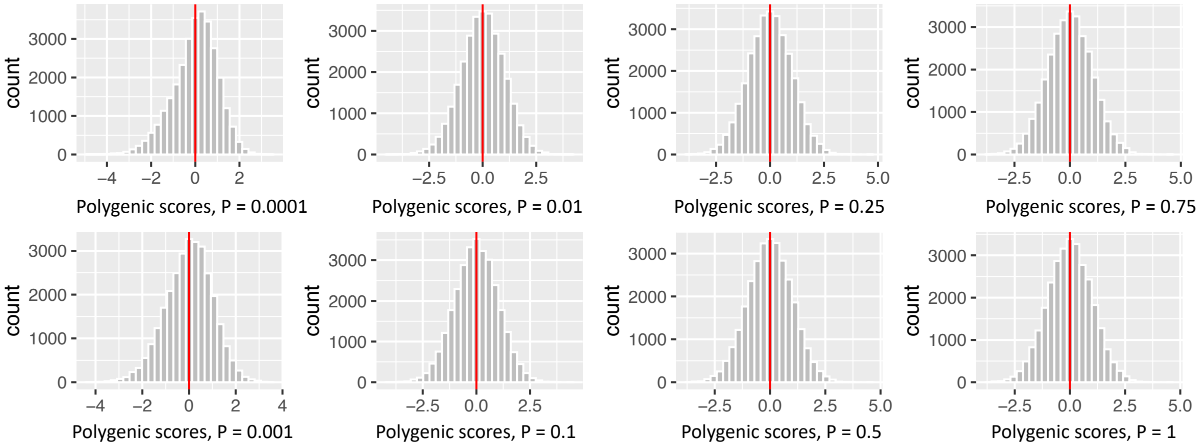

**Figure S3. Polygenic risk scores.** Frequency histograms of standardized schizophrenia polygenic risk scores in the UK Biobank at all $P_{SNP}$ value thresholds.

##### 2.2 Regional MRI phenotypes across all $P_{\text{SNP}}$ value thresholds

As outlined in the methods section, we performed a two-step analysis approach to identify the optimal $P_{\text{SNP}}$ value threshold for PRS construction for regional cortical analysis. More specifically, we first associated PRS with global cortical phenotypes and identified the most predictive $P_{\text{SNP}}$ value threshold for each phenotype ($P_{\text{SNP}}$ global). Regional cortical analyses were then performed using the most predictive $\text{P}_{\text{SNP}}$ value threshold for each phenotype. As a sensitivity test, we additionally repeated the analysis using PRS based on all other $P_{\text{SNP}}$ value thresholds. The results are shown in **Fig. S4 – S12**. For each phenotype we show the strength and direction of association (*t*-value) between all PRS and regions (A) and a Spearman's correlation matrix for each pair of PRS (B). The strength of association between schizophrenia PRS and regional MRI phenotypes was largely conserved across all $P_{\text{SNP}}$ value thresholds and was highly correlated.

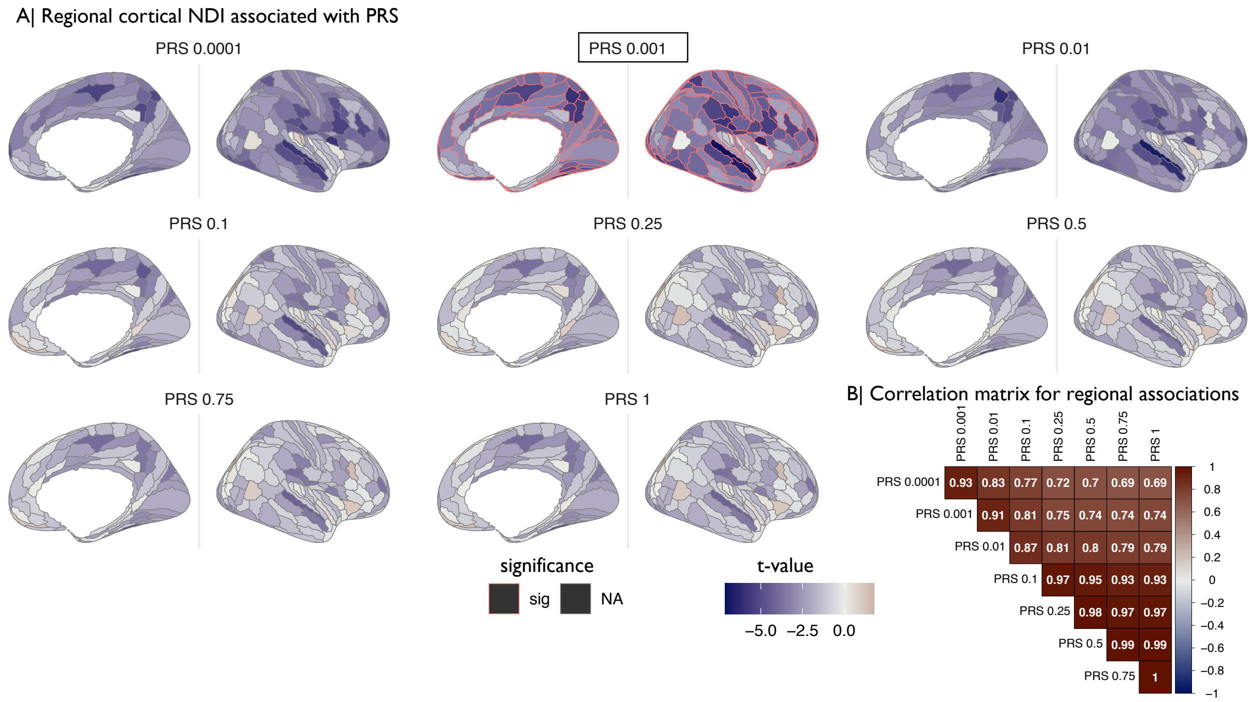

**Figure S4. Regional associations between NDI and all eight PRS based on different $P_{SNP}$ inclusion thresholds.** (A) Cortical t-maps representing strength of association between schizophrenia PRS and regional MRI phenotypes; shades of blue indicate negative associations and shades of red indicate positive associations. The PRS that was used for the main analysis is indicated with a black box and regions where the effect of the main PRS is statistically significant at FDR = 5% are outlined in red. (B) Matrix of Spearman's correlation for each pair of eight PRS.

Shades of blue indicate significant negative correlations and shades of red indicate significant positive correlations.

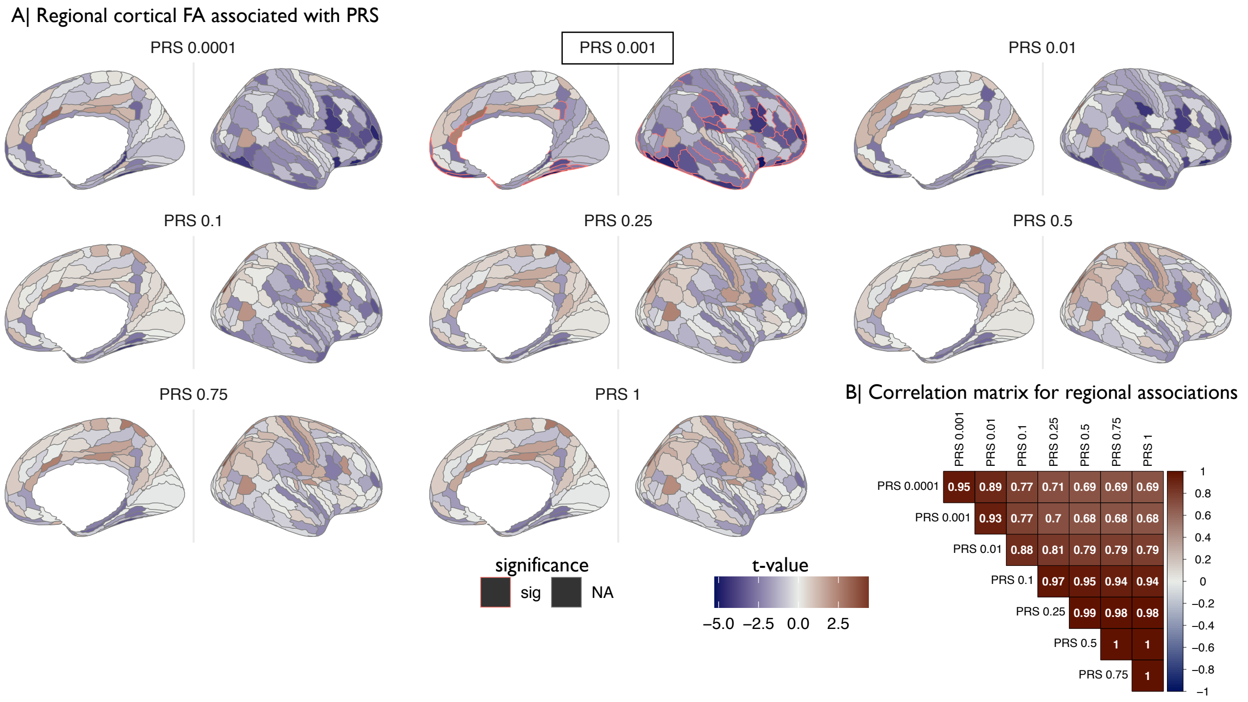

**Figure S5. Regional associations between FA and all eight PRS based on different $P_{SNP}$ inclusion thresholds.** (A) Cortical *t*-maps representing strength of association between schizophrenia PRS and regional MRI phenotypes; shades of blue indicate negative associations and shades of red indicate positive associations. The PRS that was used for the main analysis is indicated with a black box and regions where the effect of the main PRS is statistically significant at FDR = 5% are outlined in red. (B) Matrix of Spearman's correlation for each pair of eight PRS.

Shades of blue indicate significant negative correlations and shades of red indicate significant positive correlations.

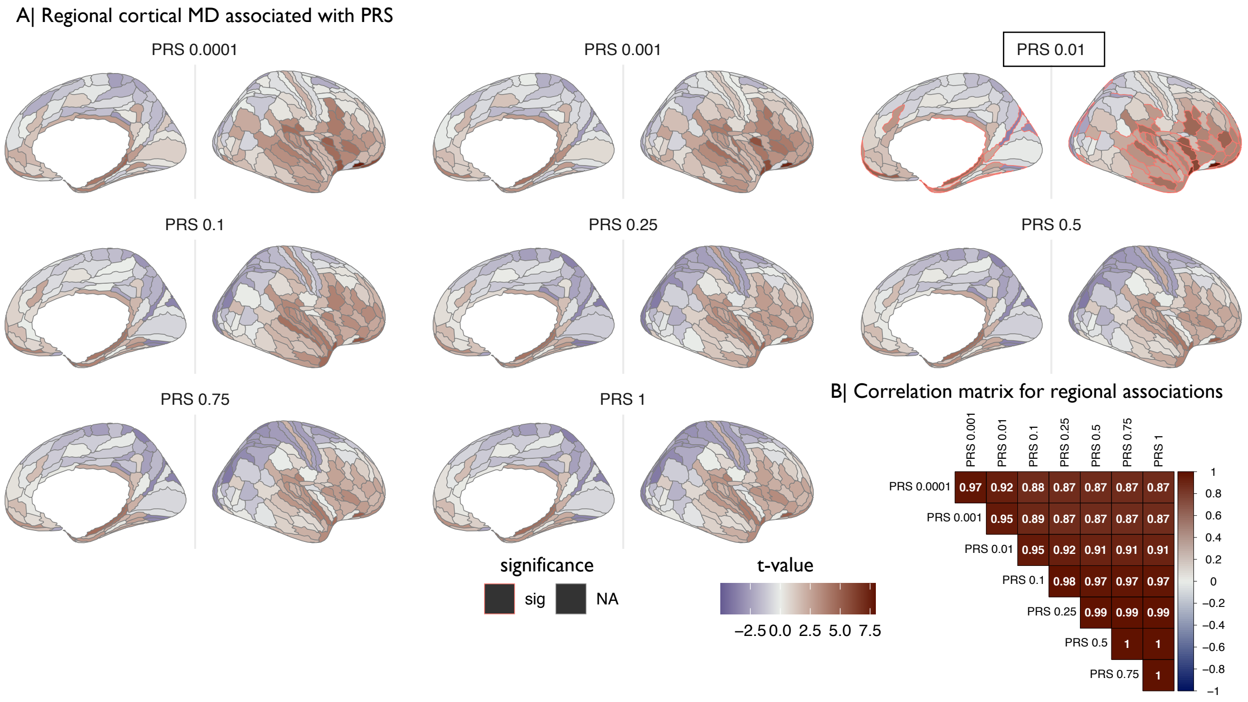

**Figure S6. Regional associations between MD and all eight PRS based on different $P_{SNP}$ inclusion thresholds.** (A) Cortical t-maps representing strength of association between schizophrenia PRS and regional MRI phenotypes; shades of blue indicate negative associations and shades of red indicate positive associations. The PRS that was used for the main analysis is indicated with a black box and regions where the effect of the main PRS is statistically significant at FDR = 5% are outlined in red. (B) Matrix of Spearman's correlation for each pair of eight PRS. Shades of blue indicate significant negative correlations and shades of red indicate significant positive correlations.

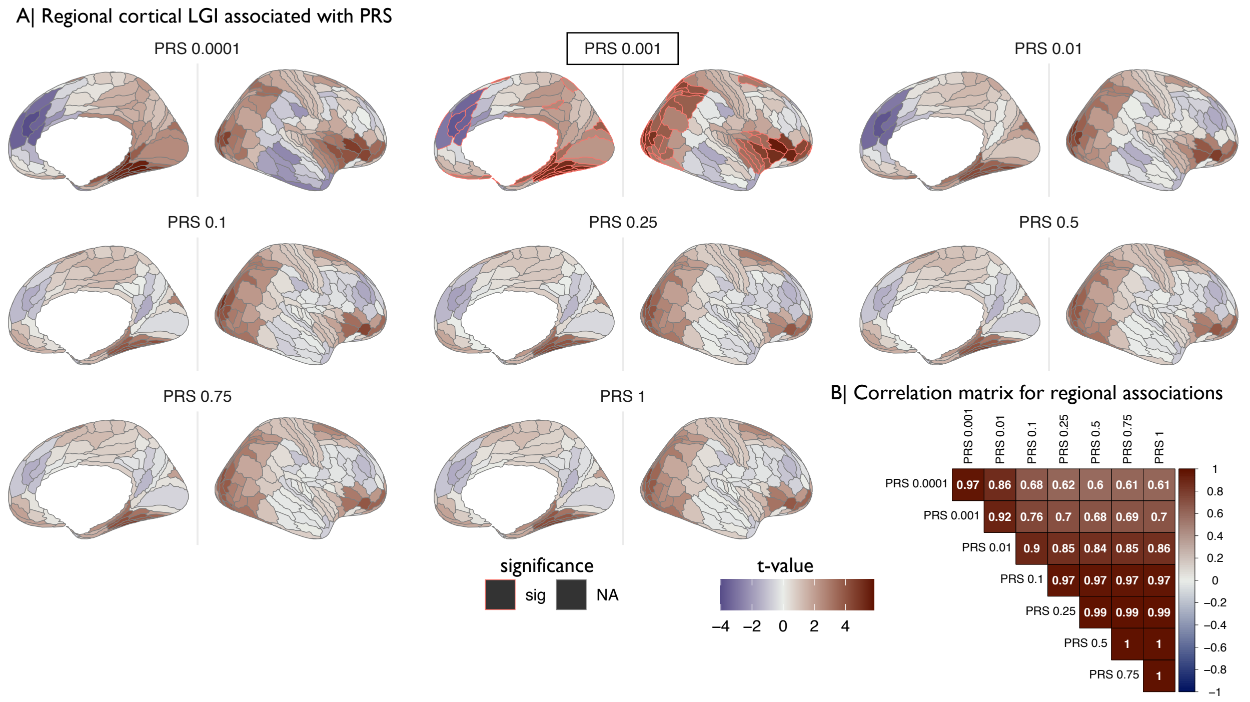

**Figure S7. Regional associations between LGI and all eight PRS based on different $P_{SNP}$ inclusion thresholds.** (A) Cortical t-maps representing strength of association between schizophrenia PRS and regional MRI phenotypes; shades of blue indicate negative associations and shades of red indicate positive associations. The PRS that was used for the main analysis is indicated with a black box and regions where the effect of the main PRS is statistically significant at FDR = 5% are outlined in red. (B) Matrix of Spearman's correlation for each pair of eight PRS. Shades of blue indicate significant negative correlations and shades of red indicate significant positive correlations.

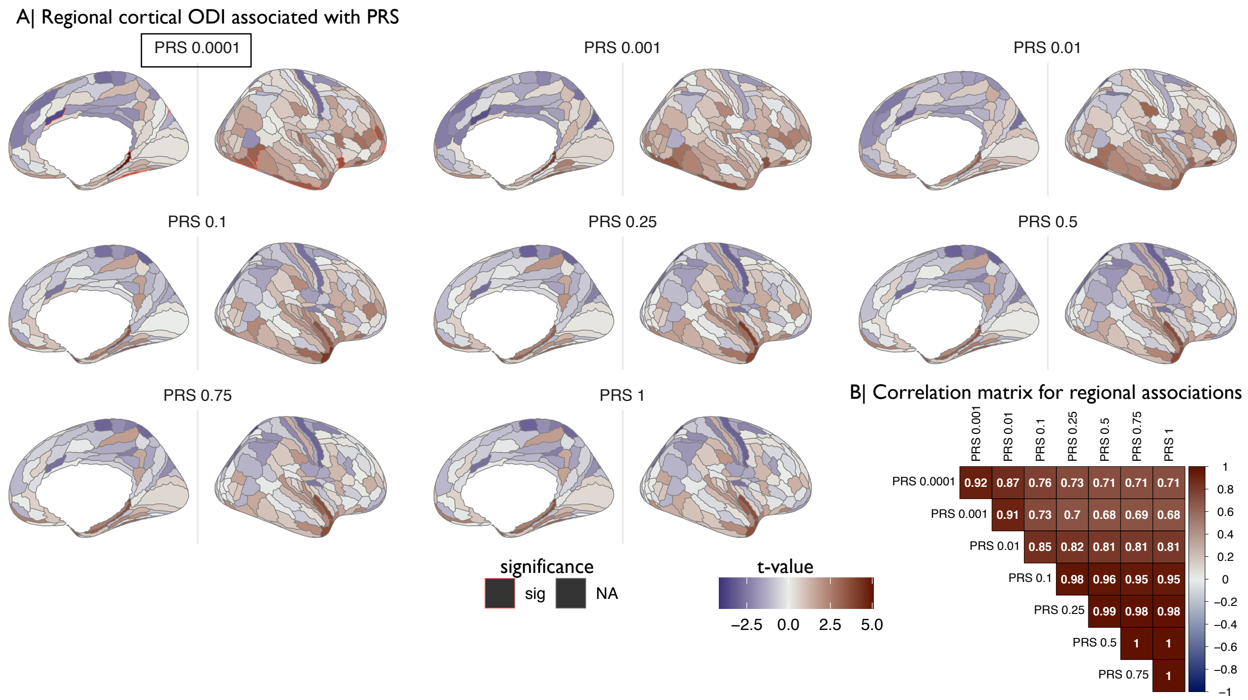

**Figure S8. Regional associations between ODI and all eight PRS based on different $P_{SNP}$ inclusion thresholds.** (A) Cortical t-maps representing strength of association between schizophrenia PRS and regional MRI phenotypes; shades of blue indicate negative associations and shades of red indicate positive associations. The PRS that was used for the main analysis is indicated with a black box and regions where the effect of the main PRS is statistically significant at FDR = 5% are outlined in red. (B) Matrix of Spearman's correlation for each pair of eight PRS. Shades of blue indicate significant negative correlations and shades of red indicate significant positive correlations.

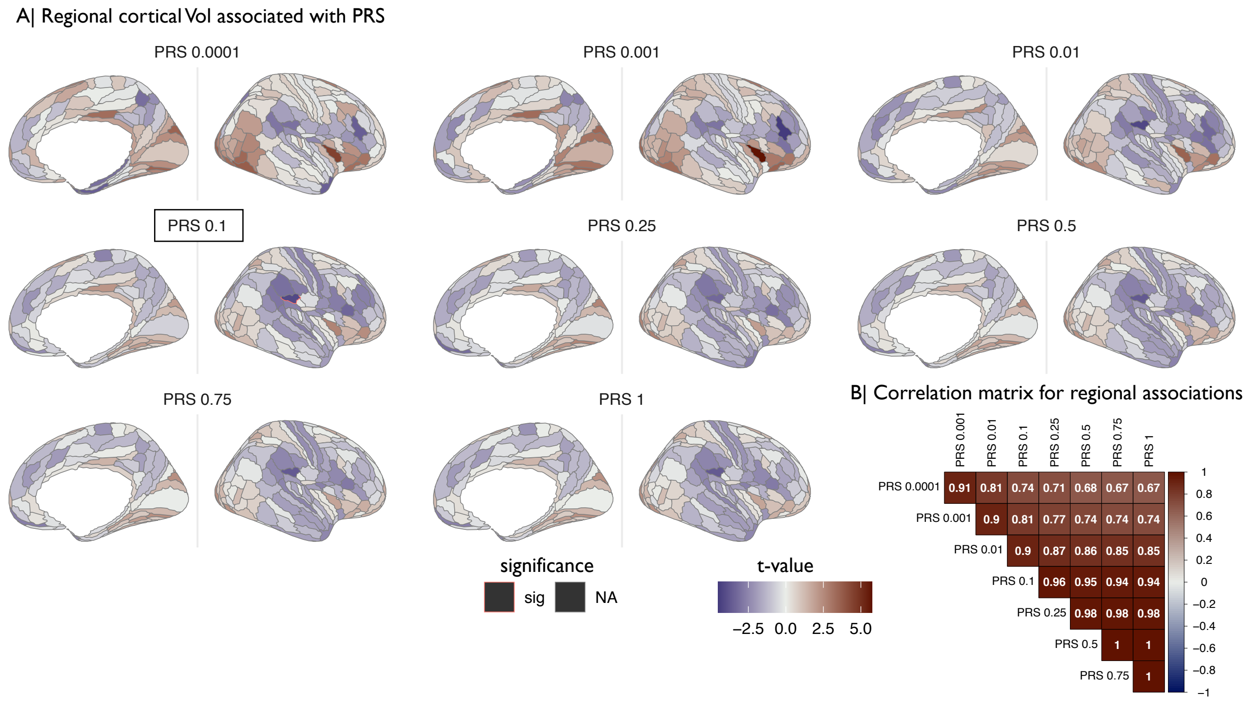

| | PRS 0.001 | PRS 0.01 | PRS 0.1 | PRS 0.25 | PRS 0.5 | PRS 0.75 | PRS 1 |
|---|---|---|---|---|---|---|---|
| PRS 0.0001 | 0.91 | 0.81 | 0.74 | 0.71 | 0.68 | 0.67 | 0.67 |
| PRS 0.001 | | 0.9 | 0.81 | 0.77 | 0.74 | 0.74 | 0.74 |
| PRS 0.01 | | | 0.9 | 0.87 | 0.86 | 0.85 | 0.85 |
| PRS 0.1 | | | | 0.96 | 0.95 | 0.94 | 0.94 |
| PRS 0.25 | | | | | 0.98 | 0.98 | 0.98 |
| PRS 0.5 | | | | | | 1 | 1 |
| PRS 0.75 | | | | | | | 1 |

**Figure S9. Regional associations between Vol and all eight PRS based on different $P_{SNP}$ inclusion thresholds.** (A) Cortical t-maps representing strength of association between schizophrenia PRS and regional MRI phenotypes; shades of blue indicate negative associations and shades of red indicate positive associations. The PRS that was used for the main analysis is indicated with a black box and regions where the effect of the main PRS is statistically significant at FDR = 5% are outlined in red. (B) Matrix of Spearman's correlation for each pair of eight PRS. Shades of blue indicate significant negative correlations and shades of red indicate significant positive correlations.

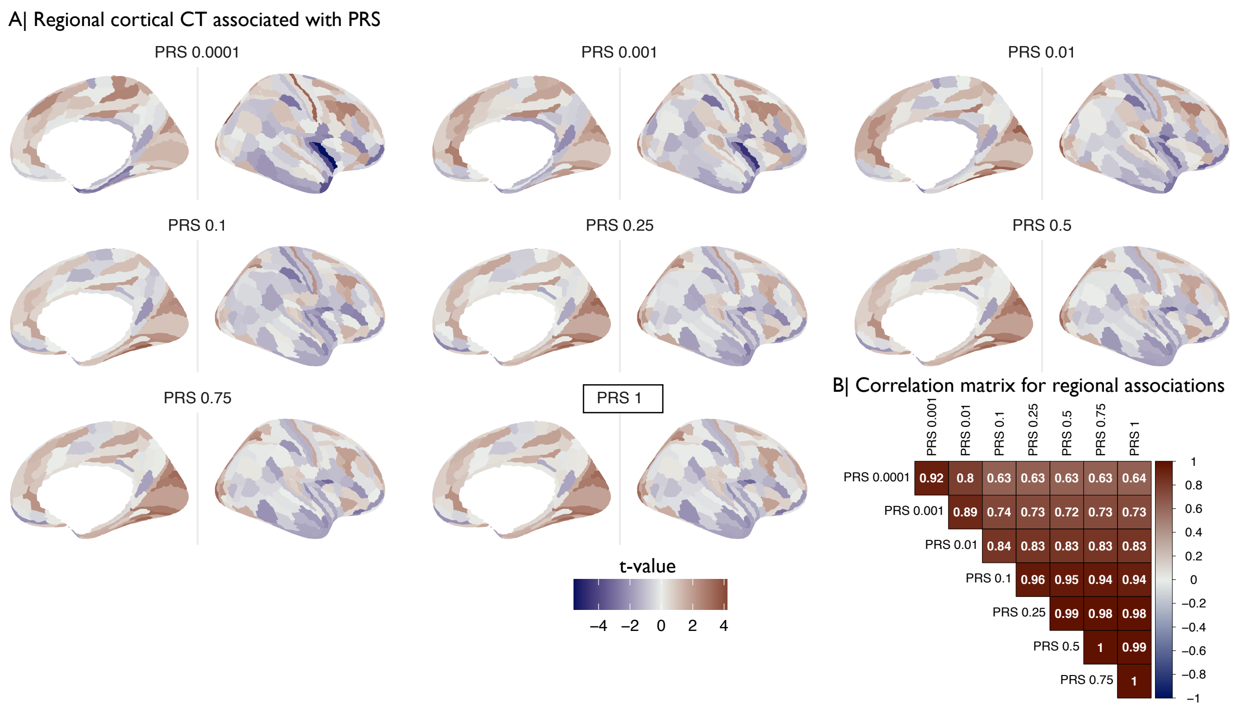

**Figure S10. Regional associations between CT and all eight PRS based on different $P_{SNP}$ inclusion thresholds.** (A) Cortical t-maps representing strength of association between schizophrenia PRS and regional MRI phenotypes; shades of blue indicate negative associations and shades of red indicate positive associations. The PRS that was used for the main analysis is indicated with a black box . For CT no region reached significance at the main PRS threshold. (B) Matrix of Spearman's correlation for each pair of eight PRS. Shades of blue indicate significant negative correlations and shades of red indicate significant positive correlations.

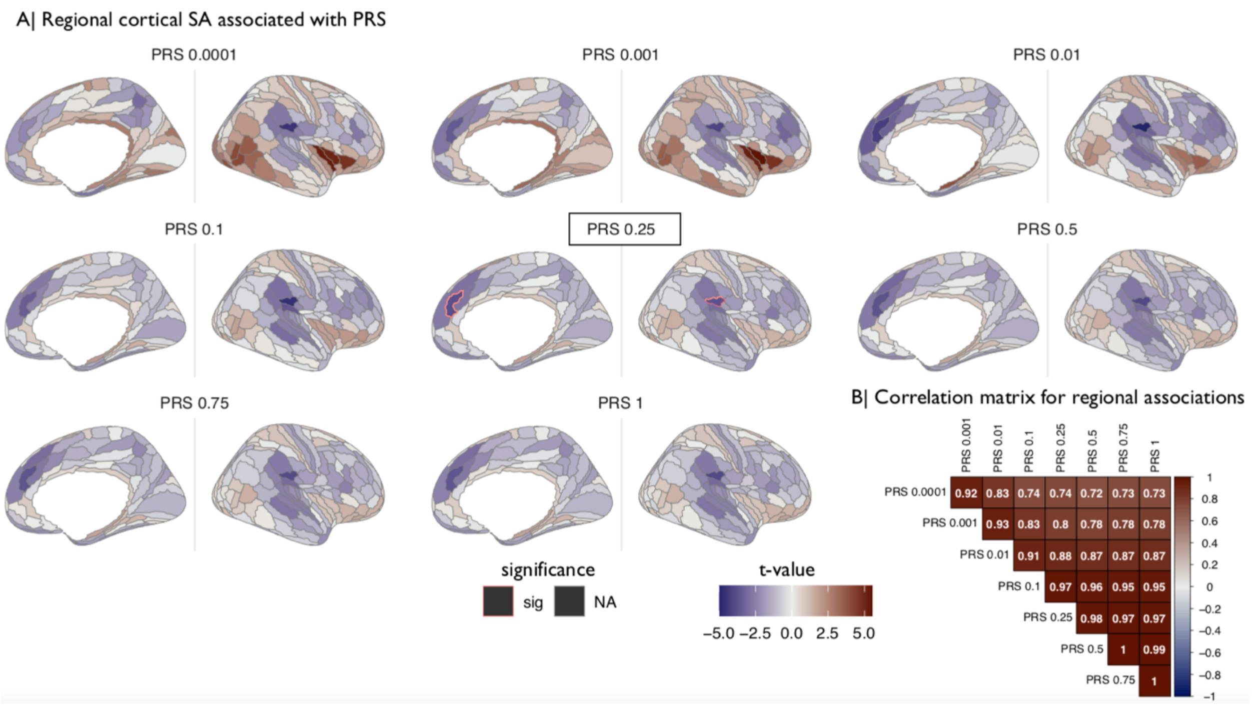

**Figure S11. Regional associations between SA and all eight PRS based on different $P_{SNP}$ inclusion thresholds.** (A) Cortical t-maps representing strength of association between schizophrenia PRS and regional MRI phenotypes; shades of blue indicate negative associations and shades of red indicate positive associations. The PRS that was used for the main analysis is indicated with a black box and regions where the effect of the main PRS is statistically significant at FDR = 5% are outlined in red. (B) Matrix of Spearman's correlation for each pair of eight PRS. Shades of blue indicate significant negative correlations and shades of red indicate significant positive correlations.

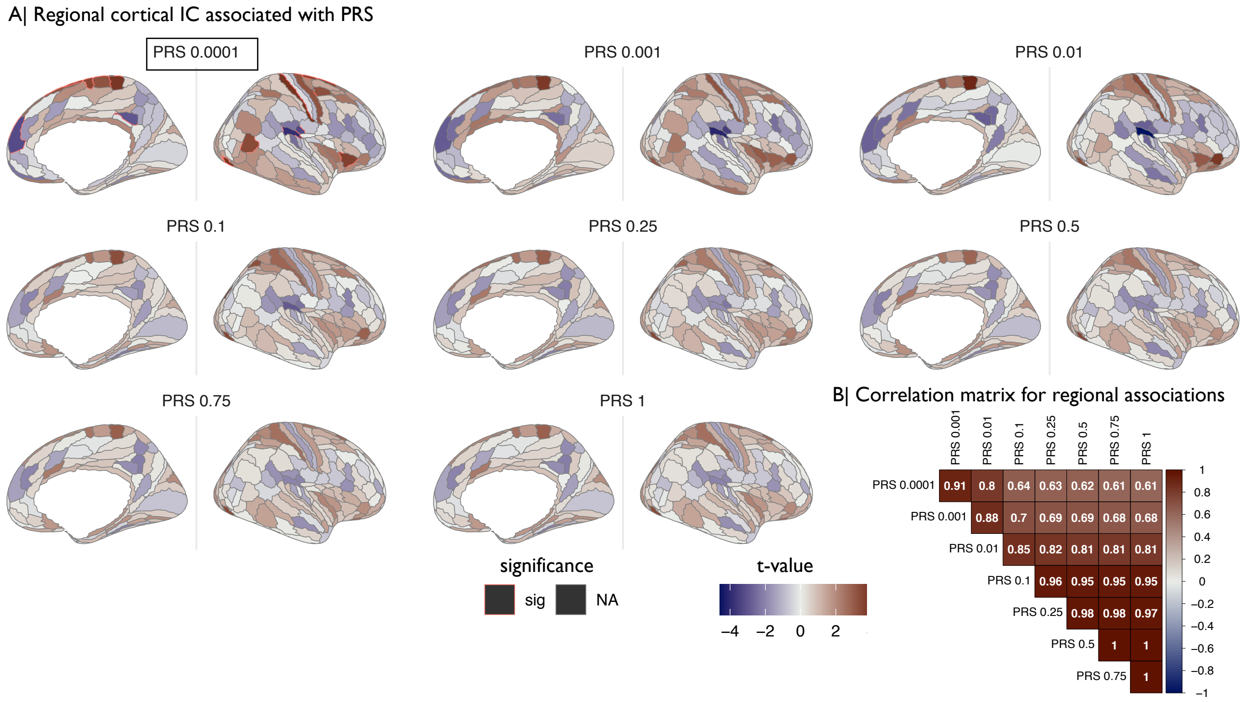

**Figure S12. Regional associations between IC and all eight PRS based on different $P_{SNP}$ inclusion thresholds.** (A) Cortical t-maps representing strength of association between schizophrenia PRS and regional MRI phenotypes; shades of blue indicate negative associations and shades of red indicate positive associations. The PRS that was used for the main analysis is indicated with a black box and regions where the effect of the main PRS is statistically significant at FDR = 5% are outlined in red. (B) Matrix of Spearman's correlation for each pair of eight PRS. Shades of blue indicate significant negative correlations and shades of red indicate significant positive correlations.

##### 2.3 Regional cortex results controlled for global measures

The mixed effect models used to investigate associations between PRS and regional cortical phenotypes included intracranial volume as a fixed effect to control for head size, as was done previously (14). As a sensitivity analysis we additionally included global measures as fixed effects to the models outlined in the main text. The strength of associations based on models including global measures as fixed effects were highly positively correlated with the strength of associations based on the models reported in the main text **(Fig. S13).**

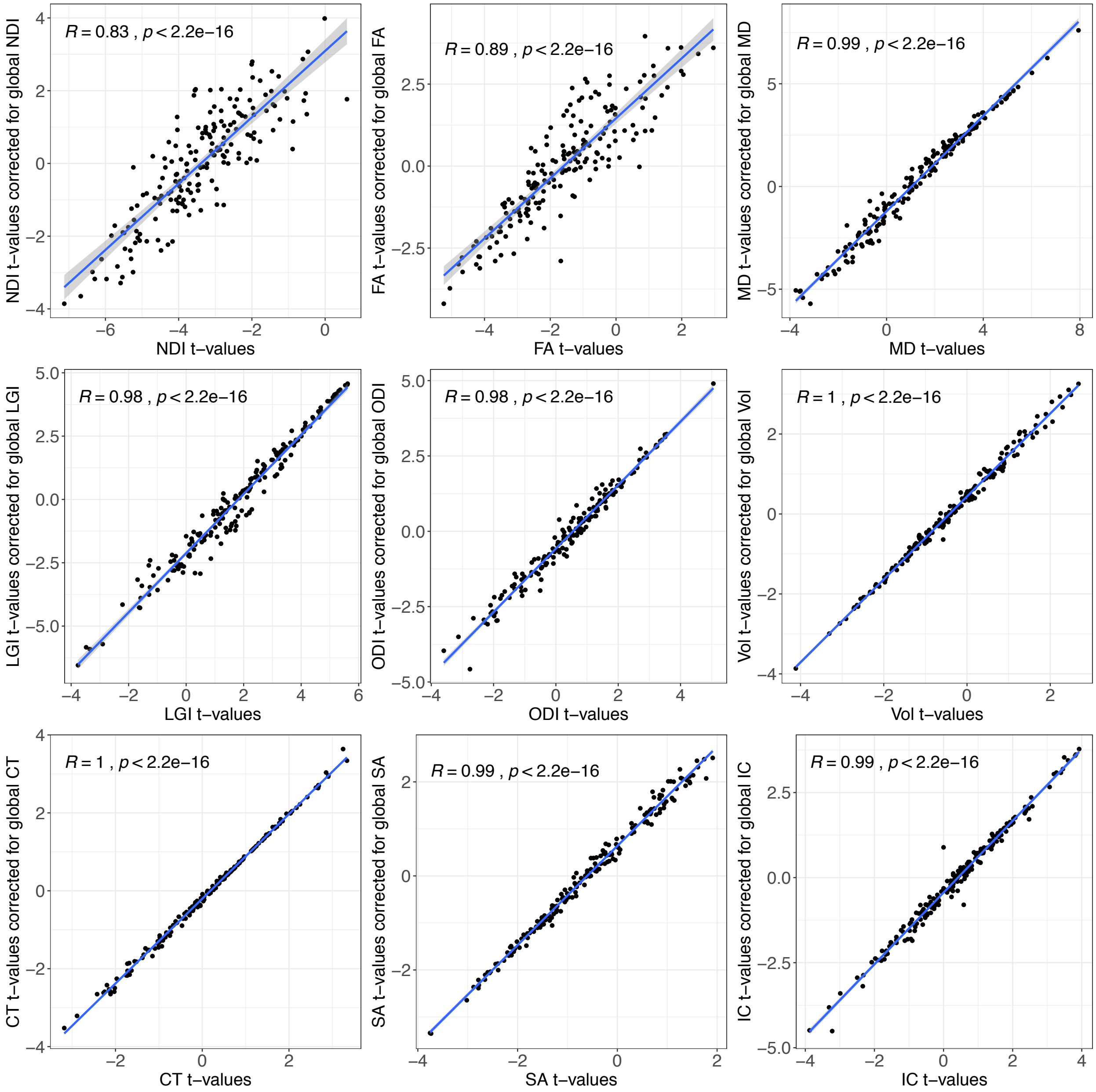

**Figure S13. Regional effects corrected for global measure.** Scatterplots showing the relationships between cortical t-maps based on linear mixed effect models controlled for global measures (y-axis) and cortical t-maps based on linear mixed effect models without global measures as a fixed effect (x-axis). Spearman's correlations and p values are shown for each imaging phenotype.

##### 2.4 Global and regional results for PRS based on European GWAS

To ensure that our findings were not biased by population stratification we repeated our analysis using polygenic risk scores based on an European GWAS (PRS EUR). PRS based on the European ancestry GWAS were normally distributed and positively correlated with PRS based on the trans-ancestry GWAS **(Fig. S14).** As shown in **Fig. S15**, global level results based on PRS EUR were overall positively correlated with the results reported in the main paper and showed a converging pattern; PRS EUR explained the highest proportion of phenotypic variance for NDI followed by FA. The proportion of variance explained by the PRS EUR was generally lower than the proportion of variance explained by the trans-ancestry PRS. This is expected, as the European ancestry GWAS was substantially smaller and therefore less powerful (10, 15). The association between PRS EUR and NDI remained significant at three $P_{SNP}$ inclusion thresholds. The association between PRS EUR and global FA no longer reached significance after correcting for multiple comparison (pFDR = 0.07 at PRS EUR  0.001) but the direction of the association remained consistent (negative association between PRS and FA). The direction of the association between PRS EUR and imaging phenotypes in this data was largely consistent with results based on the trans-ancestry PRS with 57 out of 72 associations showing the same direction of effect. The direction of the association between PRS EUR and imaging phenotypes changed for CT at three thresholds, for IC at 6 thresholds, for MD at three thresholds, for ODI at two thresholds and for SA at one threshold. We note that the direction of effect only changed for phenotypes and PRS $P_{SNP}$ value thresholds that showed non-significant and weak associations in our main results with $-.00006 \leq \beta \leq .0048$. We additionally repeated the analysis at regional level using the most predictive score from the global level analysis. Cortical t-maps based on PRS EUR were highly correlated with cortical t-maps reported in the main text **(Fig. S16).** In short, global and regional results based on PRS EUR were highly correlated with the results reported in the main paper. These findings confirm that our results based on the trans-ancestry GWAS are not driven by population stratification.

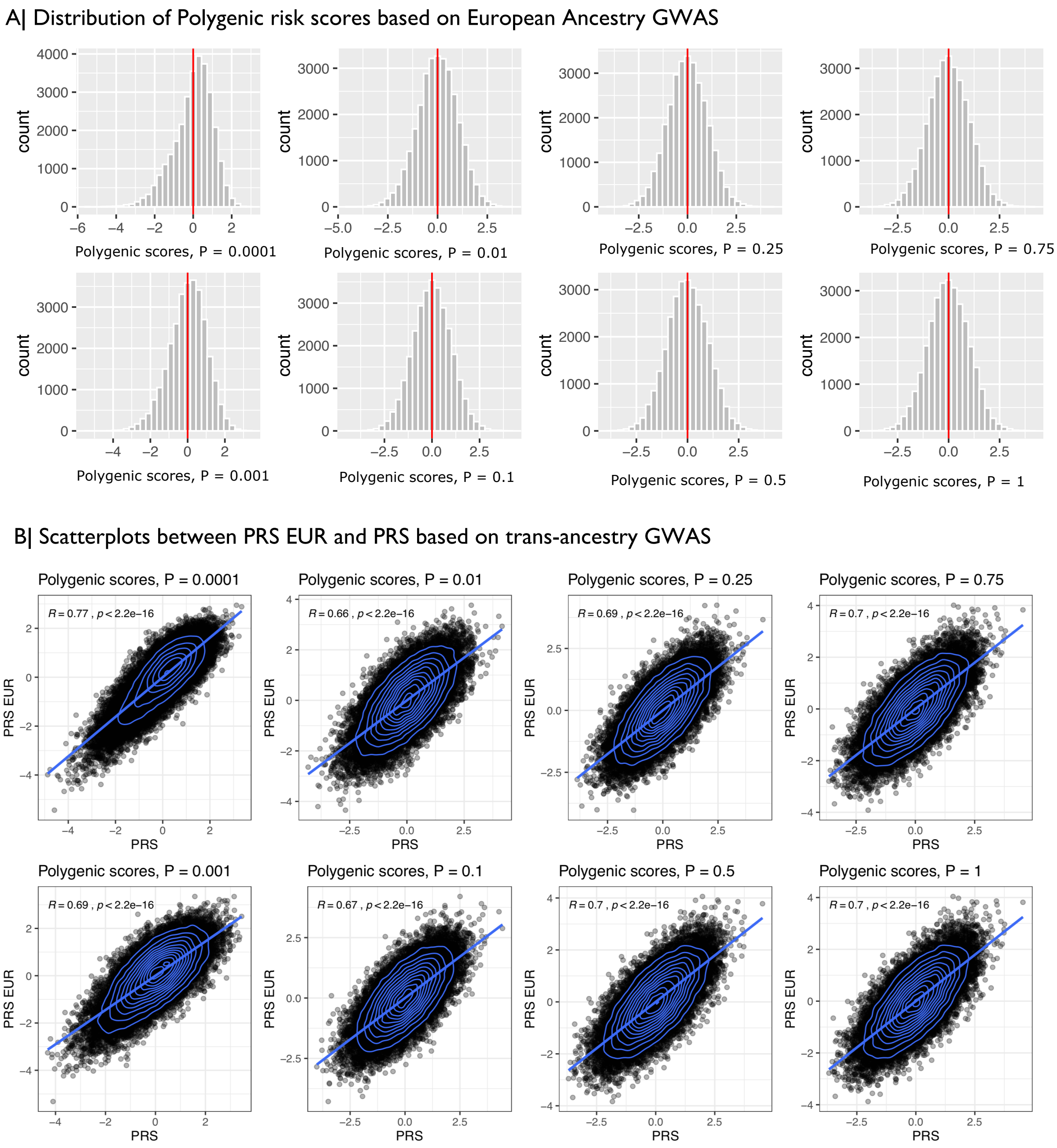

**Figure S14. Polygenic risk scores based on European ancestry GWAS.** (A) Frequency histograms of standardized schizophrenia polygenic risk scores in the UK Biobank at all $P_{SNP}$ value thresholds. (B) Scatterplots showing correlations between PRS EUR (y-axis) and PRS based on the trans-ancestry GWAS (x-axis). Spearman's correlations and p values are shown for each $P_{SNP}$ value threshold.

A| Correlation between t-values based on PRS EUR and trans-ancestry PRS

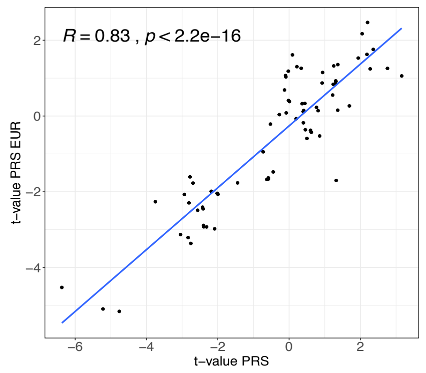

B| Global level results trans-ancestry PRS and PRS EUR

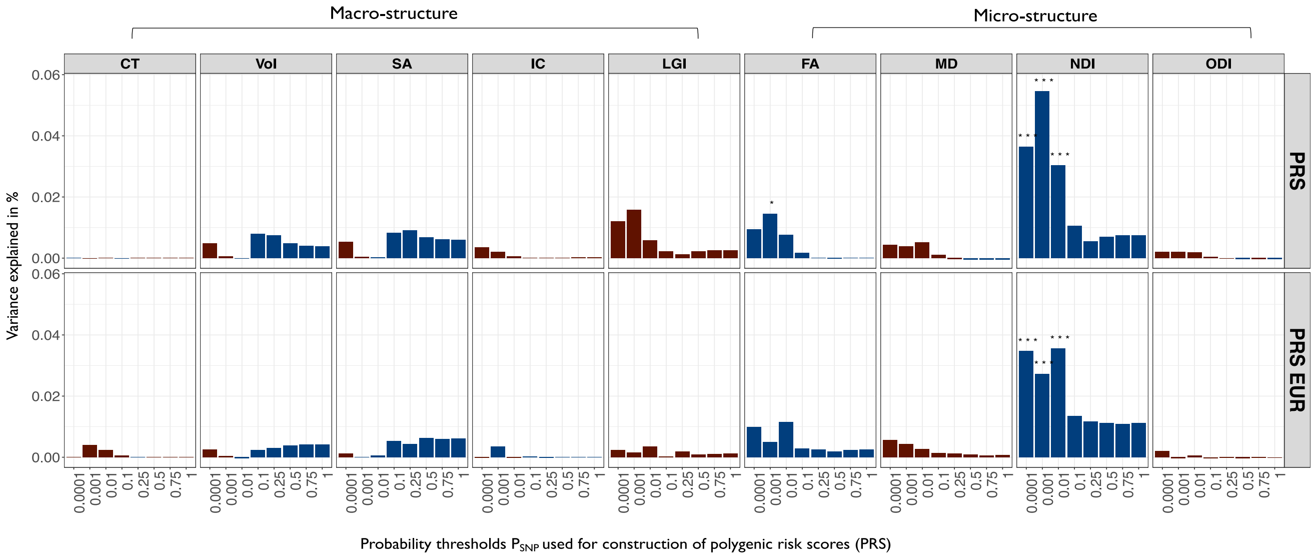

**Figure S15. Comparison between global results based on European ancestry PRS and trans-ancestry PRS*.*** (A) Scatterplot showing Spearman's correlation of t-values based on PRS EUR (y-axis) and trans-ancestry PRS (x-axis). (B) The top panel shows the global level results based on the trans-ancestry PRS reported in the main results. The lower panel shows the results based on the European ancestry PRS. Barcharts of variance explained by schizophrenia PRS ($R^2$, y-axis) constructed at each of eight probability thresholds (0.0001 ≥ $P_{SNP}$ ≤ 1, x-axis) for each of nine global mean cortical metrics: CT, cortical thickness; Vol, grey matter volume; SA surface area; IC intrinsic curvature; LGI local gyrification index; FA fractional anisotropy; MD mean diffusivity; NDI neurite density index; ODI orientation dispersion index. Blue bars indicate negative associations and red bars positive associations; asterisks indicate P-values for association after FDR correction: * P ≤ 0.05, ** P ≤ 0.01, *** P ≤ 0.001.

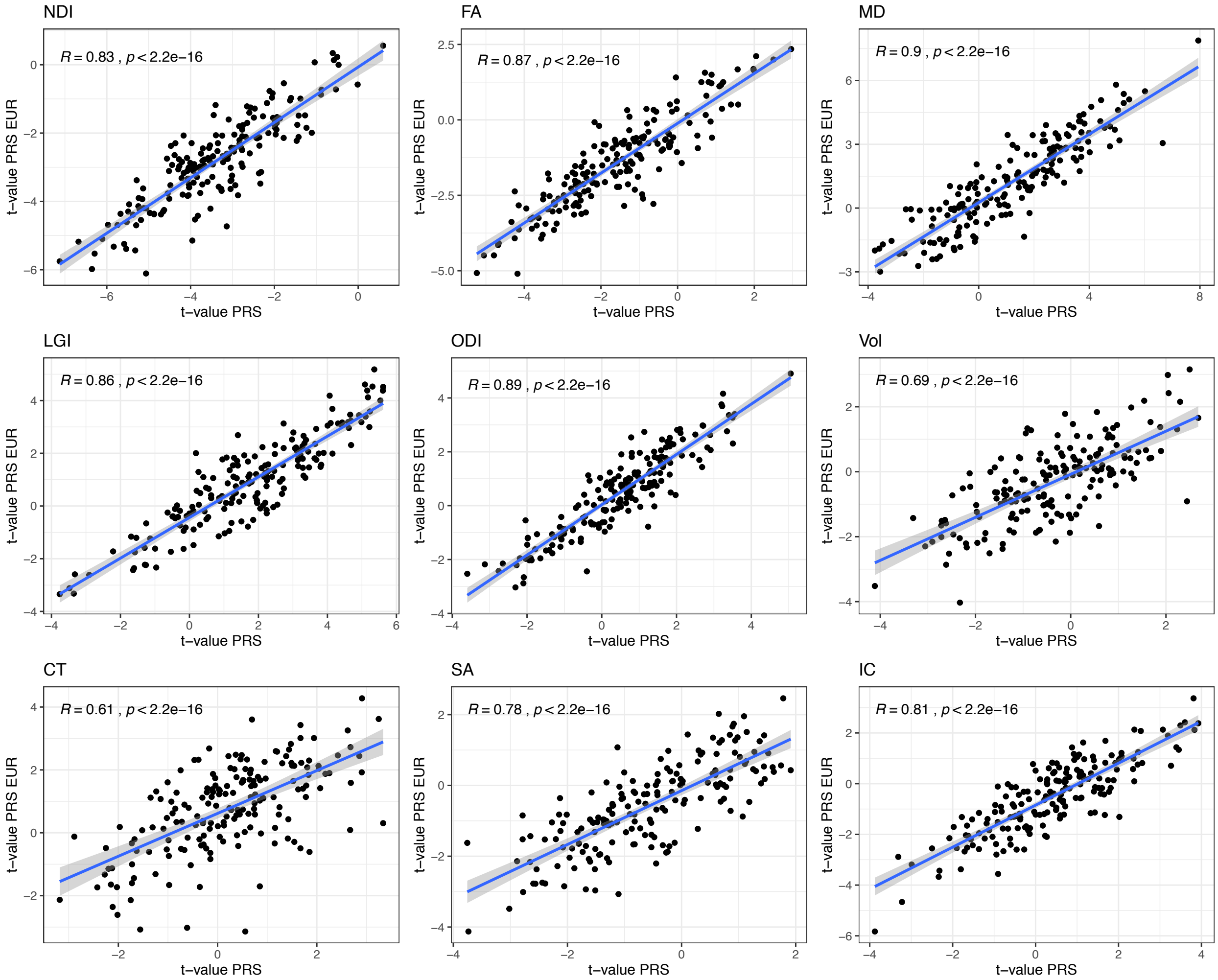

**Figure S16. Correlation between regional results based on PRS EUR and trans-ancestry PRS.** Scatterplots showing Spearman's correlation between t-values based on European PRS (y-axis) and trans-ancestry PRS (x-axis). Spearman's correlations and p-values are shown for each phenotype.

**2.5 Regional MRI phenotypes (cortical)**

At a regional cortical level, we identified significant associations between schizophrenia PRS and eight out of nine metrics in at least one cortical region with the exception being cortical thickness. The top ten strongest associations for NDI are reported in the main text. For FA the top ten strongest associations were negatively associated with the PRS-SCZ and found within ventral visual stream, insular and frontal opercular cortex, orbital and polar frontal cortex, MT+ Complex and Neighbouring Visual Areas, dorsolateral prefrontal cortex, premotor cortex and lateral temporal cortex. The top ten strongest associations for MD were positive and within orbital and polar frontal cortex, insular and frontal opercular cortex, anterior cingulate and medial prefrontal cortex, early auditory cortex and inferior frontal cortex. For LGI the top ten strongest associations were positive and within ventral visual stream, insular and frontal opercular cortex, inferior frontal cortex, medial temporal cortex, early visual cortex and medial temporal cortex. For ODI nine out of the top ten were positive and within medial temporal cortex, insular and frontal opercular cortex, orbital and polar frontal cortex, ventral stream visual, orbital and polar frontal cortex, lateral temporal cortex, dorsolateral prefrontal cortex, MT+ Complex and Neighbouring Visual Areas. One region showed a negative association and was within the anterior cingulate and medial prefrontal cortex. For Vol, we only identified one significant negative association within the early auditory cortex. The two significant associations for SA were negative within anterior cingulate and medial prefrontal cortex and early auditory cortex. Finally, eight out of the top ten strongest association for IC were positive and within early somatosensory and motor cortex, sensori-motor associated paracentral lobular and mid cingulate cortex, ventral stream visual, insular and frontal opercular cortex, sensori-motor associated paracentral lobular and mid cingulate cortex, sensory "bridge" regions of the temporal-parietal-occipital junction and the premotor cortex. The two negative associations were found in the posterior opercular cortex and the early auditory cortex **(Table S15)**.

##### 2.6 Subcortex

Within the subcortex, schizophrenia PRS explained the highest proportion of structural variance in NDI within the thalamus. However, we identified additional significant associations within all other phenotypes for at least two subcortical structures **(Fig. S17)**. Accumbens was the only subcortical structure that was not associated with schizophrenia PRS in any phenotype. We identified significant positive associations with schizophrenia PRS for MD of amygdala, caudate, hippocampus, pallidum and putamen with highest explained variance for the putamen. Additionally, we reported negative associations between schizophrenia PRS and hippocampal volume and positive associations for volume of caudate and putamen. FA and ODI showed less significant associations with two negative associations between schizophrenia PRS and FA of putamen and thalamus and significant positive associations for ODI of pallidum and thalamus **(Table S17)**.

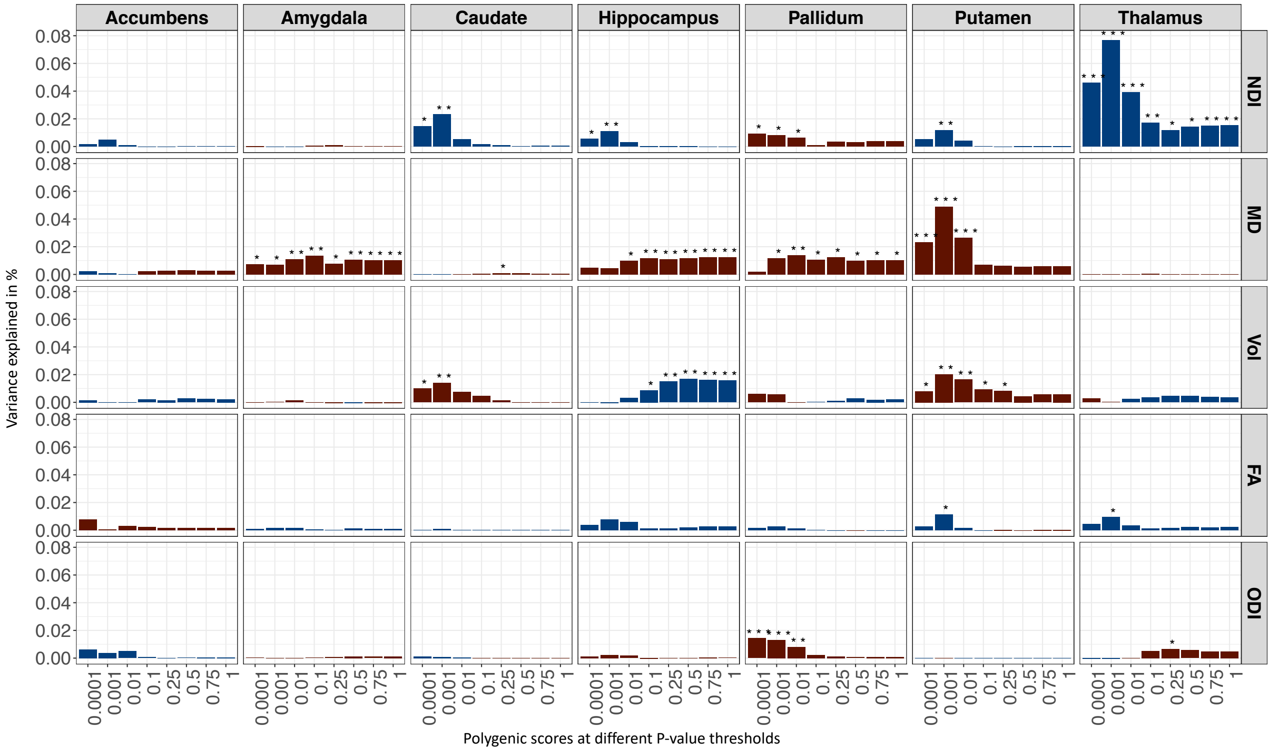

**Figure S17. Subcortical associations between polygenic risk score for schizophrenia**

**and neurite density index, mean diffusivity, volume, fractional anisotropy and orientation dispersion index of accumbens, amygdala, caudate, hippocampus, pallidum, putamen and thalamus**. Shown is the explained variance R2 (Y-axis) by the polygenic risk score at all eight P-value thresholds (X-axis). Blue bars indicate negative associations and red bars positive associations. Asterisk above bars represent P-values after FDR correction ($* \leq 0.05$, $** \leq 0.01$, $*** \leq 0.001$).

##### 2.7 Overlapping significant regions

We counted the number of overlapping significant regions for each pair of phenotypes **(Fig. S18)**. For example, 63 regions that were significantly associated with NDI also reached significance using MD. Cortical Thickness is not represented as we did not identify any significant associations between polygenic risk scores and regional cortical thickness.

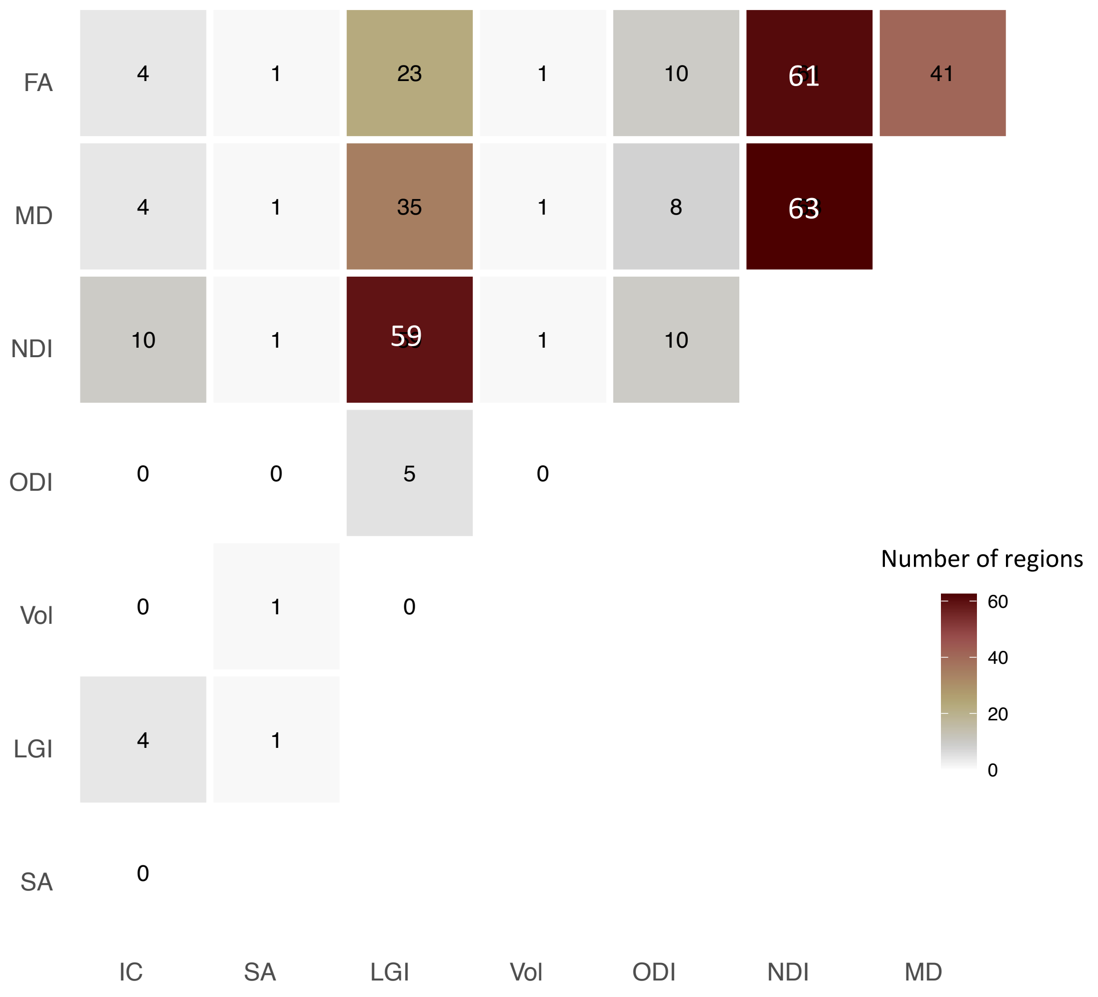

**Figure S18. Overlapping significant regions of association between PRS and each of seven cortical MRI phenotypes.**

##### 2.8 White matter tracts ODI

We investigated associations between PRS and 15 white matter tracts measured using NDI, FA, MD and ODI. In comparison to all other phenotypes ODI showed a smaller number of significant associations and a lower explained phenotypic variance. The strongest most robust associations were between PRS and ODI of uncinate fasciculus **(Fig. S19)**.

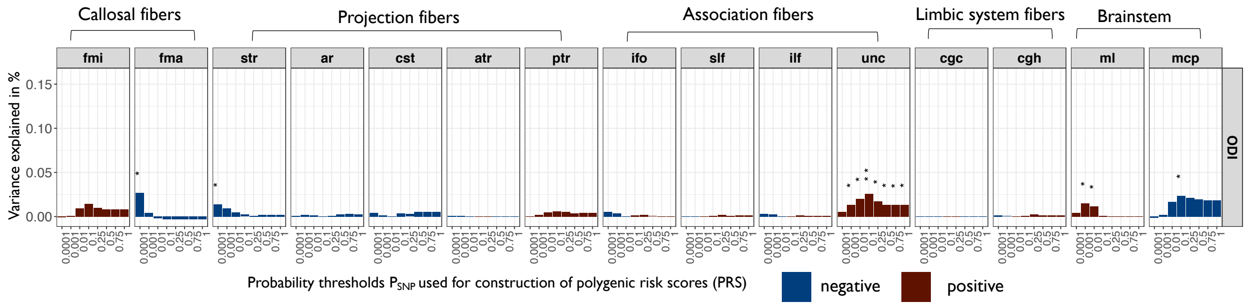

**Figure S19. Association between PRS and ODI of white matter tracts.** Barcharts of variance explained by schizophrenia PRS ($R^2$, *y*-axis) constructed at each of eight probability thresholds ($0.0001 \geq P_{SNP} \leq 1$, *x*-axis) for orientation dispersion index (ODI) measured at 15 major white matter tracts: mcp middle cerebellar peduncle; ml medial lemniscus; cst corticospinal tract; ar acoustic radiation; atr anterior thalamic radiation; str superior thalamic radiation; pts posterior thalamic radiation; slf superior longitudinal fasciculus; ilf inferior longitudinal fasciculus; ifo inferior fronto-occipital fasciculus; unc uncinate fasciculus; cgc cingulate gyrus part of cingulum; cgh parahippocampal part of cingulum; fmi forceps minor; fma forceps major. Blue bars indicate negative associations and red bars positive associations; asterisks indicate *P*-values for association after FDR correction: * $P \leq 0.05$, ** $P \leq 0.01$, *** $P \leq 0.001$.

#### 1.9 Mendelian randomization

We generated four plots to visually inspect the MR analyses for NDI of thalamus -> Schizophrenia **(Figure. S20)**.

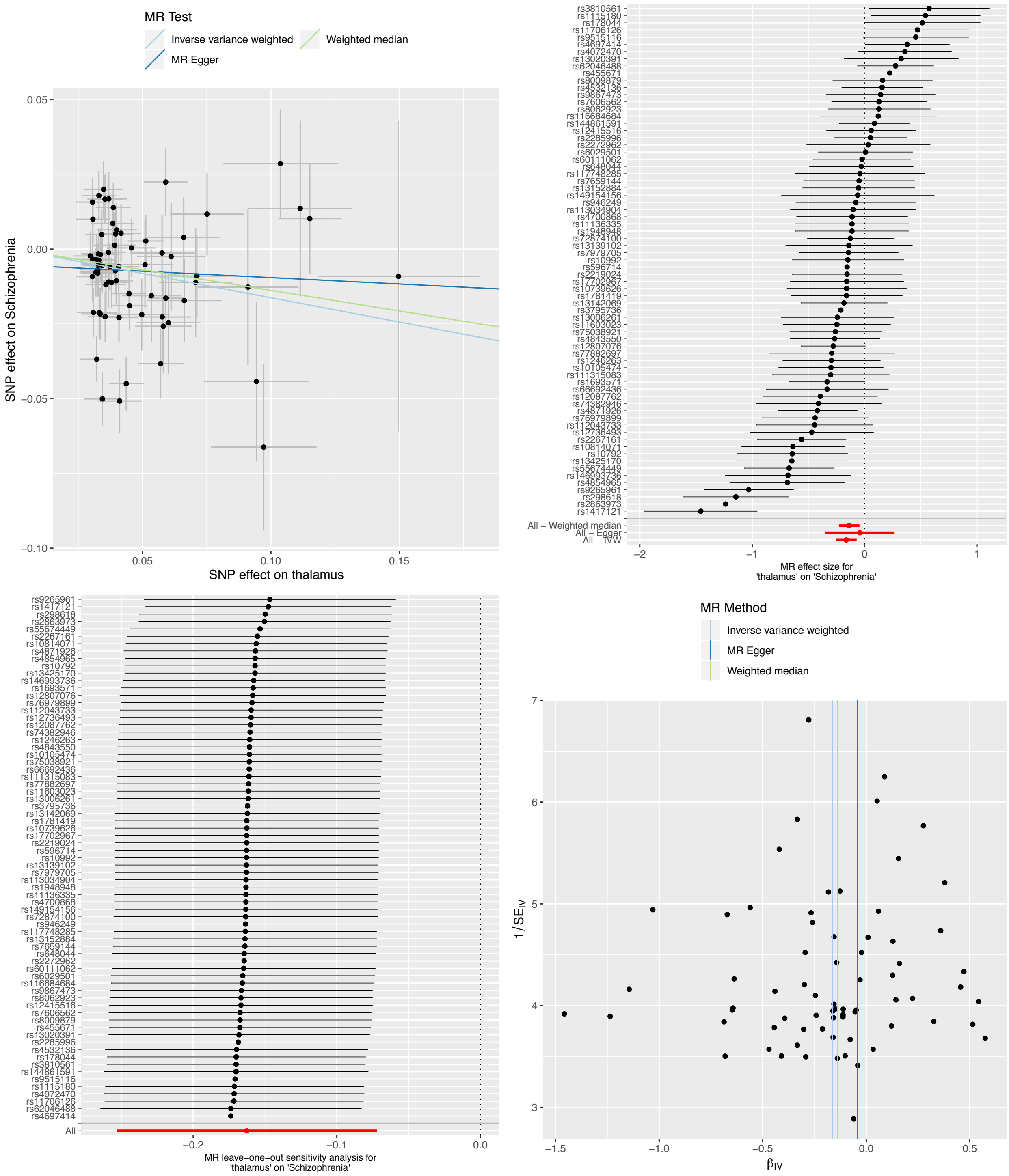

**Figure S20. Mendelian randomization analysis.** Testing the causal effect of NDI of thalamus on schizophrenia. Shown are a scatterplot (top left), forest plot (top right), leave-one-out plot (bottom left) and a funnel plot (bottom right).
